# Sex- and age-related cardiac remodelling and its association with risk factors – Results from Cardiovascular Magnetic Resonance Imaging in the German National Cohort (NAKO)

**DOI:** 10.64898/2026.03.31.26349814

**Authors:** Martyna Flis, Christopher Schuppert, Peter M. Full, Juliane Maushagen, Robin T. Schirrmeister, Marcus Dörr, Jan Gröschel, Thomas Keil, Michael Leitzmann, Wolfgang Lieb, Fiona Niedermayer, Karen Steindorf, Marco Reisert, Fabian Bamberg, Jeanette Schulz-Menger, Christopher L. Schlett, Susanne Rospleszcz

## Abstract

**Background:** The postmenopausal period is associated with a more adverse cardiometabolic risk factor profile as well as unfavourable cardiac remodelling patterns. However, it remains unclear whether and how the associations between risk factors and cardiac remodelling differ before and after menopause and in the corresponding age groups in men.

**Methods:** We used cross-sectional data from the baseline examination of the population-based German National Cohort (NAKO, age range 19-74 years). Cardiovascular resonance imaging (CMR) was performed on 3T MRI, and morphofunctional data of both ventricles were derived from standard short-axis cine balanced steady-state free precession. Associations between cardiometabolic risk factors and cardiac parameters were evaluated using adjusted multivariable linear regression, stratified by menopausal status in women and age group (<50 / ≥50 years) in men.

**Results:** The final sample comprised 20,152 participants (40% women; mean age 47±12 years) from the NAKO MRI subsample. Cardiometabolic risk factor profiles differed across the stratified groups, with higher systolic blood pressure and less favourable lipid profiles in older participants. Ventricular volumes declined and concentric remodelling increased with age in both sexes, with a steeper age-related pattern observed in women than in men. Higher BMI in women was associated with higher left ventricular concentricity index (LVCI) in postmenopausal than in premenopausal women (0.097 vs. 0.047; p for difference = 0.016). Associations between triglycerides and ventricular volumes were strongest in premenopausal women and significantly stronger than in men younger than 50 years (e.g., right ventricular end-diastolic volume (RVEDV): -0.173 vs. -0.064, p for difference < 0.001). Sleep problems were more strongly associated with cardiac parameters in men, with significant sex differences in older men compared with postmenopausal women (e.g. left ventricular end-diastolic volume (LVEDV): -0.105 vs. 0.043, p for difference = 0.023).

**Conclusions:** Less favourable cardiac remodelling observed in postmenopausal women appeared to be associated with a higher burden of cardiometabolic risk factors rather than stronger associations between these risk factors and cardiac structure. Several associations showed sex- and age-specific patterns, including Body Mass Index (BMI), triglyceride levels, and sleep problems. These findings highlight the importance of controlling cardiometabolic risk factors across adulthood, and raising awareness for sex-specific differences.

## Introduction

Cardiac remodelling reflects the heart’s structural and functional adaptation to pressure and volume overload, resulting from a complex interplay of cardiac stressors and compensatory mechanisms. In the left ventricle (LV), structural changes such as concentric remodelling, increased wall thickness, and higher ventricular mass are among the features of adverse remodelling associated with heart failure development (1, 2). These processes exhibit sex-specific differences. For example, with aging women show a more rapid increase in LV wall thickness and tend to demonstrate more concentric hypertrophy compared to men (3, 4). The postmenopausal period has been recognized as a time of accelerated cardiovascular aging (5), during which changes in hormonal homeostasis, metabolic function and vascular plasticity contribute to the elevated risk of cardiometabolic disease (6). Besides the established role of LV remodelling in cardiovascular disease (CVD), morphology and function of the right ventricle (RV) are important determinants of cardiovascular outcomes. For example, RV hypertrophy is associated with the risk of clinical heart failure or cardiovascular death in the general population (7). However, historically, research on the RV has lagged behind that of the LV, and sex- and age-related patterns of RV remodelling are less well described (8). This is partly due to the complex morphology of the RV, which is challenging to assess with standard clinical imaging such as echocardiography (9, 10).

Modifiable risk factors account for majority of the global cardiovascular disease burden and remain key targets for prevention across the life course (11). Many cardiometabolic risk factors are known to change with aging and across the menopausal transition (5). Previous large-scale cohort studies have also demonstrated that several common cardiovascular risk factors, particularly smoking and diabetes, confer disproportionately greater relative risks of CVD in women (12). These differences in clinical risk raise the question whether the associations of risk factors with subclinical LV and RV remodelling differ between pre- and postmenopausal women in the general population, or how these associations compare with those observed in men of similar ages.

Cardiac magnetic resonance imaging (CMR) provides the most accurate non-invasive assessment of cardiac structure and function, allowing quantification of LV and RV morphology and function (13). The current work is based on the imaging substudy of the German National Cohort (NAKO), a large population-based study from Germany (14), which offers an unprecedented opportunity to study CMR across the adult age range (15).

In this analysis, we aimed to 1) describe the levels of traditional metabolic and lifestyle CVD risk factors in the population-based NAKO study for an age range covering the pre- and postmenopausal period, 2) describe the sex-specific subclinical remodelling patterns of both cardiac ventricles, as derived by CMR, 3) assess whether the associations between these risk factors and ventricular remodelling phenotypes differ between premenopausal women, postmenopausal women and the corresponding age groups in men.

## Methods

### Study population

We used data from the Magnetic Resonance Imaging (MRI) subsample of the NAKO study. The NAKO study is a population-based, prospective cohort study conducted across 18 study centers in Germany. The baseline examination took place between 2014 and 2019 and involved 205,415 participants aged 19-74 years (14). The baseline protocol included standardized interviews, biomedical examinations, questionnaires, and biosamples collection. Whole-body MRI scans were performed at five study centres on a 3.0-T MR scanner (Magnetom Skyra; Siemens Healthcare, Erlangen, Germany), using an identical hardware and software setup. Within on average 30 days after the baseline examination, 30,868 participants underwent the MRI examination (16).

### Outcome assessment: Cardiac Magnetic Resonance

CMR parameters were obtained from standard short-axis cine balanced steady-state free precession images (15, 17). Measures included parameters of morphology and function for the left and right ventricles, comprising left ventricular ejection fraction (LVEF), end-diastolic volume (LVEDV), end-systolic volume (LVESV), stroke volume (LVSV), and cardiac output (LVCO), as well as right ventricular ejection fraction (RVEF), end-diastolic volume (RVEDV), end-systolic volume (RVESV), stroke volume (RVSV), and cardiac output (RVCO). Additional left ventricular measures included concentricity index (LVCI), wall thickness (LVWT), and end-diastolic mass (LVM). LVCI was calculated as LVM/LVEDV. For further analysis, volumes, cardiac outputs (CO), wall thickness, and mass were indexed to height raised to an allometric power of 1.7 (18).

### Assessment of covariates and risk factors

Participants underwent standardized physical examinations, interviews, and questionnaires at the study centres. Information on previous cardiovascular disease (CVD), including myocardial infarction, coronary heart disease, heart failure, arrhythmias, and peripheral arterial disease, as well as the use of antihypertensive, antidiabetic, and lipid-lowering medications, was self-reported. Body mass index (BMI) was calculated as weight divided by height squared (kg/m²). Blood pressure was measured using standardized protocols and calibrated devices. Lipid profile, including low-density lipoprotein cholesterol (LDL-C), high-density lipoprotein cholesterol (HDL-C), and triglycerides, as well as haemoglobin A1c (HbA1c), was assessed using standard laboratory procedures (14).

Information on lifestyle factors including smoking status, alcohol use, physical activity and sleep was self-reported. Smoking status was classified as never smoker, ex-smoker, and current smoker. Pack-years were defined as the number of packs smoked per day multiplied by the duration of smoking in years. Alcohol intake was calculated in units, with one unit being equal to 8 g of ethanol. Physical activity was defined as moderate-to-vigorous physical activity during leisure time and was quantified in metabolic equivalent hours (MET-hours/week). Sleep quality was assessed based on sleep length, and a binary variable indicating the presence or absence of sleep problems. Sleep length was further categorized into short (<6.5 h), normal (6.5–8.5 h), and long (>8.5 h) (14).

### Definition of postmenopause and age groups

Postmenopause was defined as one of the following: no menstrual bleeding for more than 6 consecutive months in women aged 45 years or older, permanent cessation of menstruation due to illness, or age greater than 60 years.

We identified the age at which the upper bound of the premenopausal age distribution (90th percentile) and the lower bound of the postmenopausal age distribution (10th percentile) converged and used this threshold to stratify men into younger (<50 years) and older (≥50 years) groups corresponding to pre- and postmenopausal women, respectively. Group comparisons were conducted between premenopausal and postmenopausal women, premenopausal women and younger men, postmenopausal women and older men, and younger versus older men.

### Statistical methods

Sample characteristics are presented as mean and standard deviation (SD) for continuous variables and as counts and percentage for categorical variables. Values more than 4.5 SD from the mean were treated as outliers and removed. Differences between groups were assessed using Welch’s *t*-test and χ²-test respectively, with *p*-values adjusted for multiple comparisons by the false discovery rate (FDR) method.

Risk factors and CMR parameters according to age were described by generalized additive models (GAMs) using thin plate regression splines as the smoothing basis. Smooth functions were visualized using GAM plots with 95% confidence intervals.

Associations between metabolic and lifestyle risk factors and CMR parameters were examined using multivariable linear regression models. The following variables were included as exposures: BMI, systolic blood pressure, diastolic blood pressure, LDL-C, triglycerides, HbA1c, smoking status, alcohol consumption, physical activity, sleep duration and sleep problems. All exposures were entered simultaneously in the same model. Models were additionally adjusted for HDL-C and use of antihypertensive, antidiabetic, and lipid-lowering medications. Estimates are reported as β-coefficients with 95% confidence intervals (CIs), with β-coefficients appropriately scaled (metabolic factors standardized to z-scores; lifestyle factors on the original scale). To account for the cumulative effects of smoking exposure, models were additionally adjusted for pack-years in a sensitivity analysis to assess the robustness of the associations.

Differences in regression coefficients between groups were assessed using a z-test, with corresponding two-sided *p*-values derived from the standard normal distribution (19). We considered *p*-values <0.05 after correction for false discovery rate (FDR) as statistically significant. All statistical analyses were done with R version 4.4.2.

## Results

### Study sample

The final analytical sample comprised 20,152 participants, after the exclusion of individuals with poor-quality CMR images or extreme outliers, those with prevalent CVD or unknown CVD status, and those with missing information on menopausal status, lifestyle, or metabolic risk factors (Supplementary Figure 1). Participants had a mean age of 47 ± 12 years, and 40% were women. Mean systolic blood pressure was 127 ± 16 mmHg and mean diastolic blood pressure was 79 ± 10 mmHg. Regarding smoking status, 51% of participants had never smoked, 31% were former smokers, and 18% were current smokers. Current smoking was most prevalent among younger men (<50 years; 21%), whereas the highest proportion of former smokers was observed among older men (41%) (Table 1).

**Table 1.**
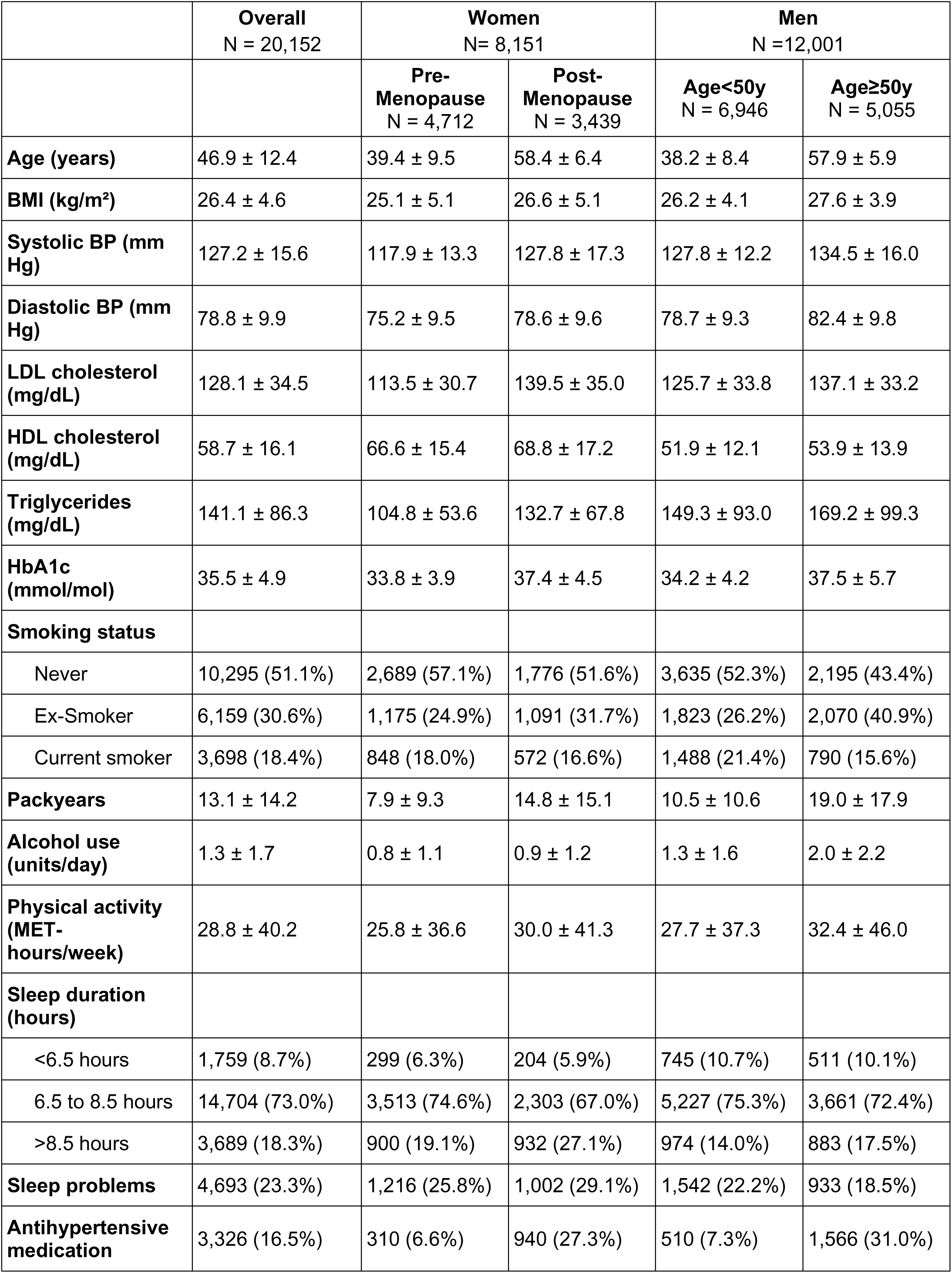
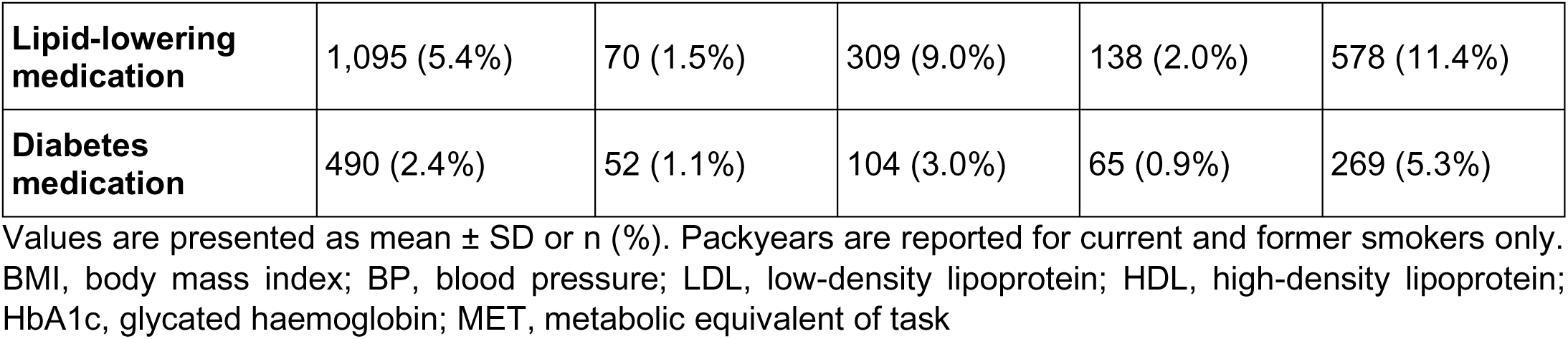
Characteristics of the study sample according to menopausal status in women, and age group in men.

In the overall sample, mean LVEF was 63.1%. Younger adults showed higher ventricular ejection fractions (EF) than older adults, and women consistently demonstrated higher LVEF than men within each age group. Mean LVSV was 35.3 ml/m^1.7^, with younger individuals again having higher stroke volumes than older individuals, and women showing lower values than men. Left ventricular mass averaged 44.6 g/m^1.7^ overall and was markedly higher in men than in women across both age groups, with older adults exhibiting greater mass than younger adults (Table 2).

**Table 2.** Cardiac parameters of the study sample, as derived by CMR, according to menopausal status in women, and age group in men.

|  | **Overall**<br>N = 20,152 | **Women**<br>N= 8,151 |  | **Men**<br>N =12,001 |  |
| --- | --- | --- | --- | --- | --- |
|  |  | **Pre-Menopause**<br>N = 4,712 | **Post-Menopause**<br>N = 3,439 | **Age<50y**<br>N = 6,946 | **Age≥50y**<br>N = 5,055 |
| **LV Ejection Fraction (%)** | 63.1 ± 5.4 | 64.5 ± 4.9 | 64.8 ± 5.2 | 62.1 ± 5.2 | 62.0 ± 5.7 |
| **LV End-diastolic Volume (ml/m<sup>1.7</sup>)** | 56.0 ± 10.5 | 55.4 ± 8.6 | 50.9 ± 8.8 | 59.6 ± 10.7 | 55.1 ± 10.9 |
| **LV End-systolic Volume (ml/m<sup>1.7</sup>)** | 20.7 ± 5.1 | 19.7 ± 4.2 | 17.9 ± 4.2 | 22.6 ± 5.2 | 20.9 ± 5.2 |
| **LV Stroke Volume (ml/m<sup>1.7</sup>)** | 35.3 ± 7.1 | 35.7 ± 6.1 | 33.0 ± 6.2 | 37.0 ± 7.3 | 34.2 ± 7.5 |
| **LV Cardiac Output (ml/min\*m<sup>1.7</sup>)** | 2,236.9 ± 571.3 | 2,258.3 ± 531.9 | 2,090.0 ± 523.5 | 2,343.4 ± 582.1 | 2,171.0 ± 592.2 |
| **RV Ejection Fraction (%)** | 55.6 ± 7.0 | 56.7 ± 6.8 | 58.5 ± 6.8 | 53.9 ± 6.7 | 55.1 ± 6.9 |
| **RV End-diastolic Volume (ml/m<sup>1.7</sup>)** | 63.1 ± 12.2 | 60.9 ± 10.2 | 55.2 ± 9.8 | 68.1 ± 12.1 | 63.6 ± 12.3 |
| **RV End-systolic Volume (ml/m<sup>1.7</sup>)** | 28.1 ± 7.4 | 26.4 ± 6.3 | 22.9 ± 5.5 | 31.4 ± 7.4 | 28.5 ± 7.0 |
| **RV Stroke Volume (ml/m<sup>1.7</sup>)** | 35.0 ± 7.7 | 34.5 ± 6.8 | 32.3 ± 7.0 | 36.7 ± 7.8 | 35.1 ± 8.2 |
| **RV Cardiac Output (ml/min\*m<sup>1.7</sup>)** | 2,219.8 ± 602.8 | 2,178.9 ± 552.0 | 2,047.5 ± 553.5 | 2,326.3 ± 612.2 | 2,229.5 ± 635.7 |
| **LV Concentricity Index (g/ml)** | 0.8 ± 0.2 | 0.7 ± 0.1 | 0.8 ± 0.1 | 0.8 ± 0.1 | 0.9 ± 0.2 |
| **LV Wall Thickness (mm/m<sup>1.7</sup>)** | 2.8 ± 0.4 | 2.6 ± 0.3 | 2.8 ± 0.4 | 2.8 ± 0.3 | 3.0 ± 0.4 |
| **LV Mass (g/m<sup>1.7</sup>)** | 44.6 ± 8.3 | 38.2 ± 5.5 | 38.8 ± 5.9 | 48.4 ± 7.0 | 49.4 ± 7.5 |

Values are presented as mean ± SD. LV, left ventricle; RV, right ventricle

### Metabolic and lifestyle risk factors levels across the age range

In women, BMI rose steadily across the age range, whereas in men, BMI was higher in early and mid-adulthood and then stabilized or slightly declined after age 60 (Figure 1). Systolic blood pressure, LDL-C, and triglyceride levels were generally lower in women than in men, but they increased more sharply in women during midlife (Figure 1). Consequently, women’s systolic blood pressure and triglyceride levels approached those of men in later life, and women’s LDL-C levels even exceeded men’s, who showed a plateau or slight decline at older ages. Physical activity and sleep duration showed a U-shaped relationship with age, with higher levels in younger and older adults and lower levels during midlife (Figure 1).

**Figure 1.**
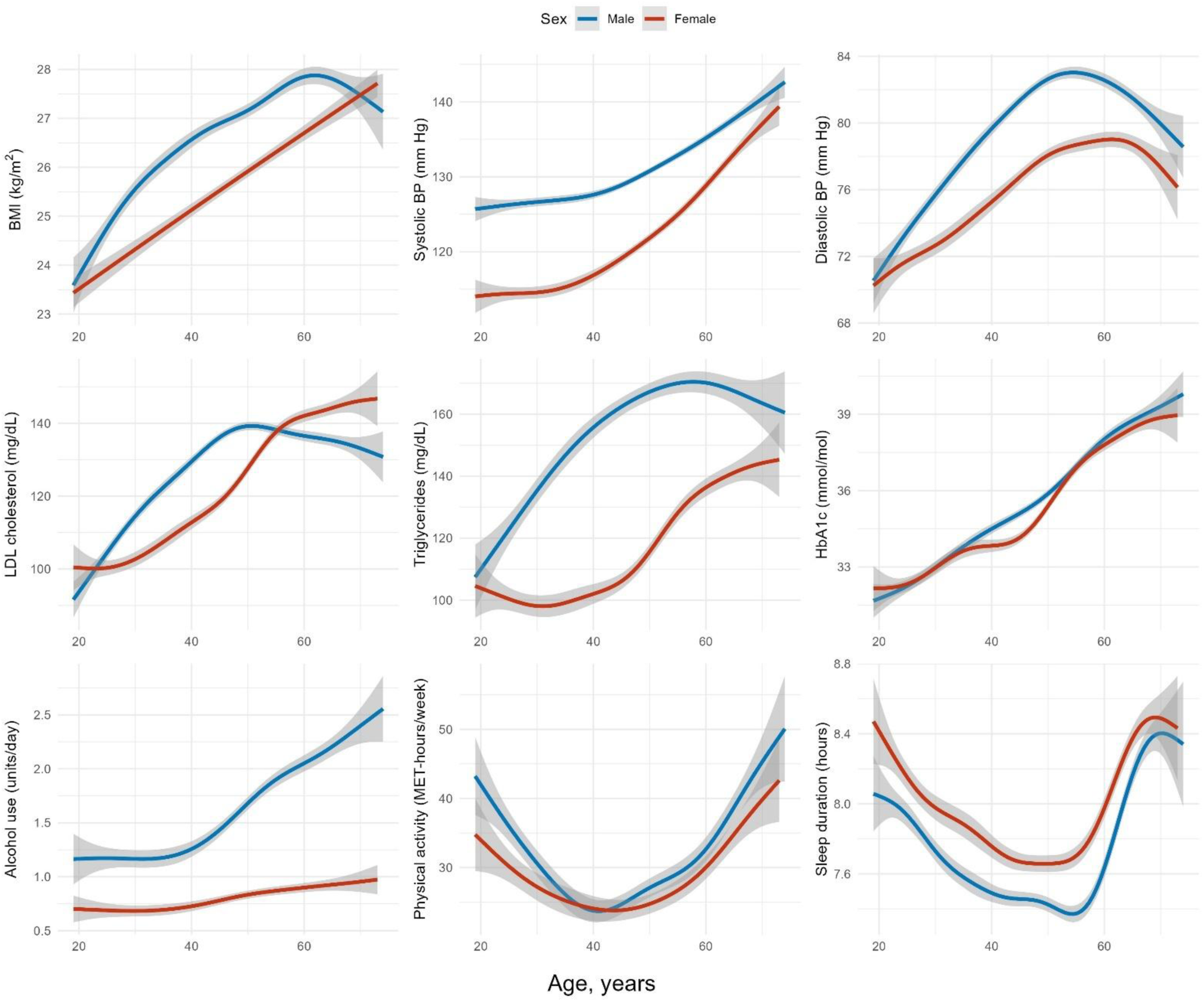
Continuous risk factors by sex and age. Figures display age (years) on the x-axis and the unadjusted GAM-estimated mean of the risk factor conditional on age on the y-axis. Grey shading indicates 95%-CIs.

### Cardiac parameters across the age range

LVEF in women increased gradually until midlife and then remained relatively stable, whereas in men it rose to a lesser extent and began to decline around the same age (Figure 2). Cardiac volumes declined with age in both sexes. In men, volumes decreased linearly across age groups, whereas in women, the decline accelerated with age, remaining stable at younger ages and becoming progressively steeper in later life (Figure 2). Structural measures, LVWT and LVCI, increased steadily with age, with a change in slope observed at midlife. After this point, the rate of increase became steeper, particularly among women.

**Figure 2.**
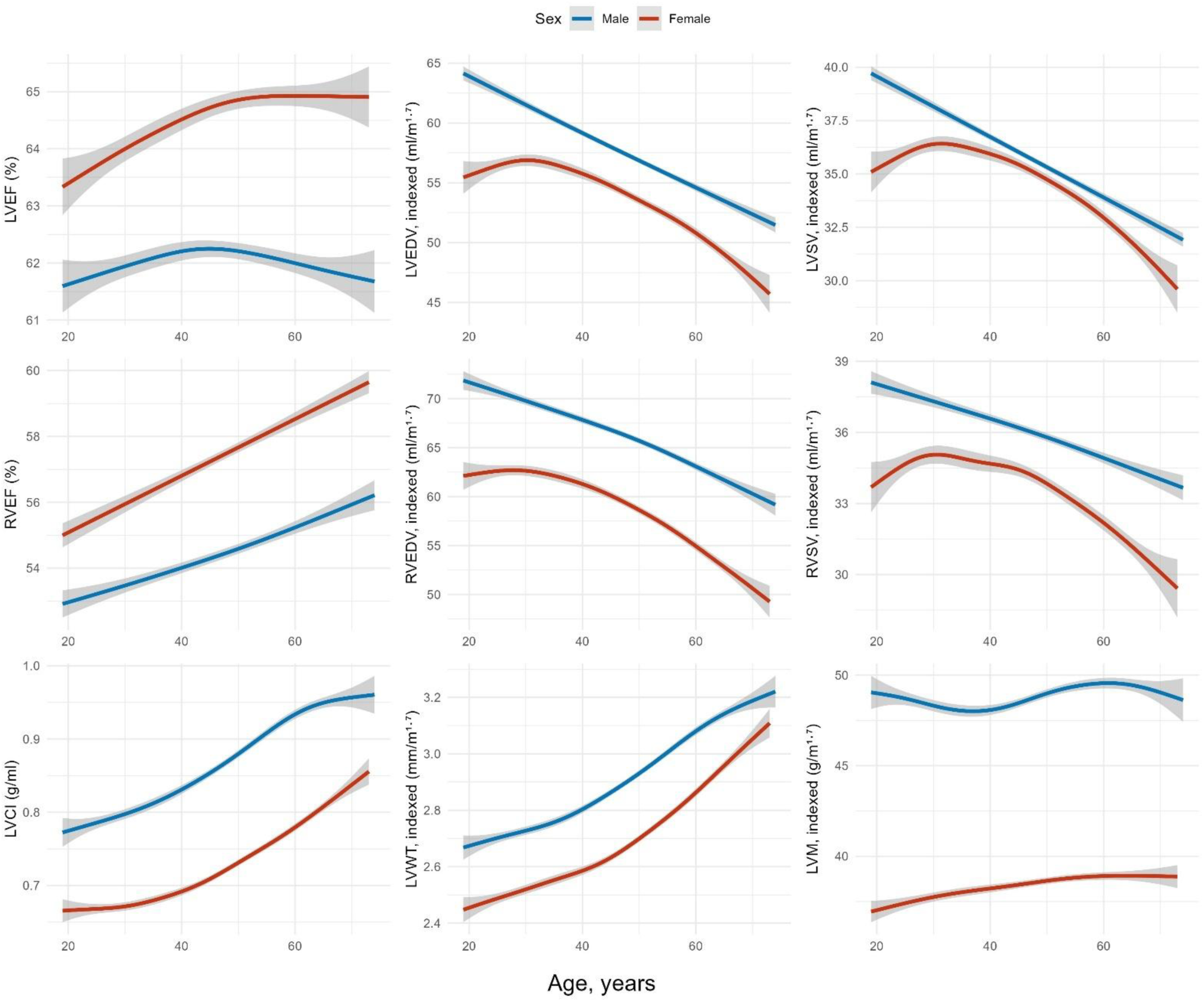
Continuous risk factors by sex and age. Figures display age (years) on the x-axis and the unadjusted GAM-estimated mean of the cardiac parameter conditional on age on the y-axis. Grey shading indicates 95%-CIs.

### Associations of metabolic risk factors with cardiac parameters

Higher BMI in women was associated with lower ejection fraction in both ventricles, with a more pronounced association for LVEF in postmenopausal than in premenopausal women (–0.090 vs –0.009; p for difference = 0.005; Supplementary Table 1; Figure 3). BMI showed positive associations with ventricular volumes, which were consistently stronger in premenopausal women (e.g., LVEDV: 0.306 vs. 0.241; p for difference= 0.006; Supplementary Table 1; Figure 3). BMI was also positively related to LVCI, LVWT, and LVM, although the association with LVCI was stronger in postmenopausal women (0.097 vs. 0.047; p for difference = 0.016, Supplementary Table 1; Figure 3).

**Figure 3.**
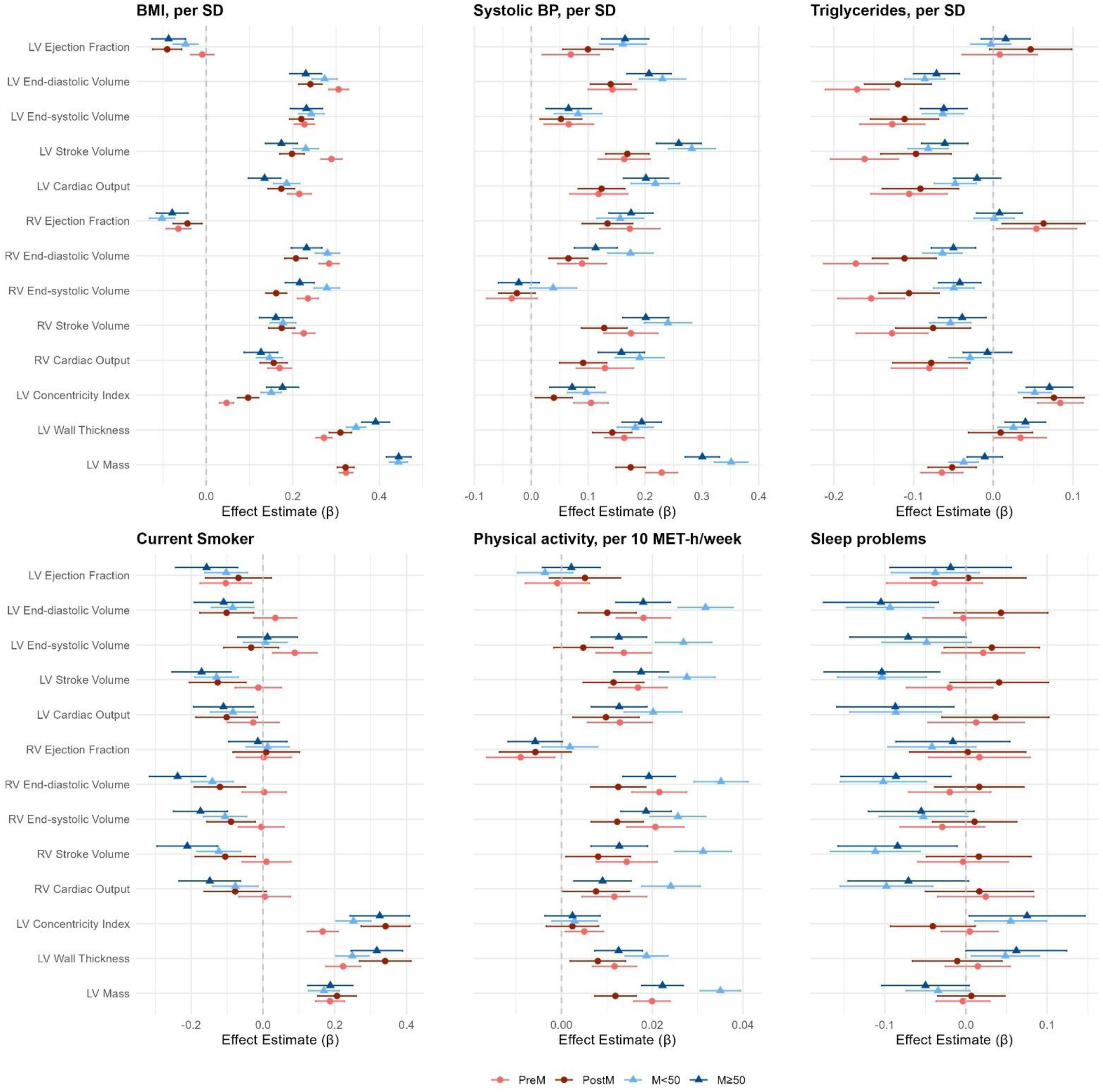
Forest plots of standardized associations between selected lifestyle and cardiometabolic risk factors and cardiovascular outcomes, stratified by sex and age/menopausal status. Estimates are shown per SD increase (or per category for categorical variables) with 95% confidence intervals, based on fully adjusted models fitted separately in each group.

In men, higher BMI was associated with lower EF in both ventricles, while all remaining cardiac measures showed positive associations. Across both age groups, BMI demonstrated stronger associations with LVCI, LVWT, and LVM in men than in women (e.g., LVM: 0.444 to 0.445 vs. 0.322 to 0.323; p for difference < 0.001, Supplementary Table 1; Figure 3).

Higher systolic blood pressure in women was associated with higher EF and larger ventricular volumes, as well as greater LVCI, LVWT, and LVM, with stronger associations observed in premenopausal than postmenopausal women (e.g., LVCI: 0.105 vs. 0.040; p for difference= 0.040, Supplementary Table 2; Figure 3). Higher diastolic blood pressure, in contrast, was linked to lower EF, smaller volumes, and lower CO in both ventricles, but to higher LVCI and LVWT.

In men, higher systolic blood pressure was associated with higher EF and volumes for both ventricles, as well as higher LV mass and concentricity. Higher diastolic BP in men was associated with lower EF, volumes and CO in both ventricles. Negative associations between diastolic BP and ventricular volumes were more pronounced in younger men (e.g., LVEDV: –0.316 vs. –0.237; p for difference = 0.043, Supplementary Table 3). Diastolic blood pressure was additionally linked to higher LVCI and LVWT, but showed a negative association with LVM. In both age groups, systolic and diastolic blood pressure showed more pronounced associations in men than in women for most cardiac parameters, except LVCI or LVWT.

Higher LDL-C levels in women were associated with lower cardiac volumes, reduced CO, and lower LVM, with no significant differences between premenopausal and postmenopausal women (Supplementary Table 4). Triglyceride levels were similarly linked to lower volumes and CO in both ventricles, with numerically stronger associations in premenopausal than postmenopausal women (e.g., RVEDV: –0.173 vs. –0.112; p for interaction = 0.155, Supplementary Table 5; Figure 3).

Notably, triglyceride associations with volumes were significantly stronger in premenopausal women than in men under 50 years (e.g., RVEDV: –0.173 vs. –0.064; p for difference < 0.001, Supplementary Table 5; Figure 3), whereas corresponding differences between younger and older men were comparatively small (e.g., RVEDV: –0.064 vs. –0.050; p for difference = 0.707, Supplementary Table 5; Figure 3).

Triglycerides were also associated with higher LVCI and LVWT in both men and women. HbA1c showed a comparable pattern to triglycerides, with associations reflecting lower cardiac volumes and CO alongside higher LVCI, LVWT, and LVM in all groups (Supplementary Table 6).

### Associations of lifestyle risk factors with cardiac parameters

Current smoking was associated with lower LVEF in both pre- and postmenopausal women. No reduction in cardiac volumes was observed in premenopausal women, whereas broader adverse associations were observed in postmenopausal women (e.g., LVEDV: 0.034 vs. –0.101; p for difference = 0.053, Supplementary Table 7; Figure 3). Smoking was also associated with higher LVCI, LVWT, and LVM in both groups, with a stronger association for LVCI in postmenopausal women (0.166 vs. 0.342; p for difference < 0.001, Supplementary Table 7; Figure 3)

In men, former smoking was associated with lower cardiac volumes and higher LVCI and LVWT across age groups. Current smoking was linked to lower LVEF, reduced biventricular volumes and cardiac output, and higher LVCI, LVWT, and LVM. Men younger than 50 years showed stronger adverse associations of former smoking with cardiac volumes than premenopausal women, along with a greater increase in LVCI (0.106 vs. -0.002; p for difference = 0.006, Supplementary Table 8). After additional adjustment for pack-years, the previously observed differences between young men and postmenopausal women relative to premenopausal women were attenuated (Supplementary table 9).

Higher alcohol consumption was associated with increased LVM in postmenopausal women, and in both age groups in men (Supplementary Table 10). Physical activity showed moderate positive associations with cardiac volumes and outputs, as well as LVWT and LVM in both premenopausal and postmenopausal women. In men, physical activity was moderately positively associated with cardiac volumes, outputs, and structure with consistently stronger associations in men younger than 50 years in both ventricles (e.g., LVM: 0.022 vs. 0.035; p for difference = 0.003, Supplementary Table 11; Figure 3).The associations between physical activity and cardiac structure and function were consistently strongest in younger men, and significantly stronger than in premenopausal women (e.g., LVM: 0.035 vs. 0.020; p for difference < 0.001, Supplementary Table 11; Figure 3).

Short sleep duration was associated with higher LVWT in both sexes, whereas long sleep duration was associated with lower LVEF, cardiac volumes, and CO. Associations of sleep duration did not differ significantly between men and women (Supplementary Table 12; Supplementary Table 13). Self-reported sleep problems were more strongly associated with cardiac parameters in men, with significant sex differences in older men compared with postmenopausal women (e.g. LVEDV: -0.105 vs. 0.043; p for difference = 0.023, Supplementary Table 14; Figure 3).

## Discussion

In this population-based study, we characterized sex-specific patterns of cardiometabolic risk factors and MRI-derived measures of cardiac structure and function across the adult age range. Triglyceride levels were most strongly associated with cardiac structural parameters in premenopausal women, whereas BMI showed a stronger relationship with adverse remodelling in postmenopausal compared with premenopausal women. In addition, sleep disturbances were more strongly associated with cardiac measures in men. With respect to menopause, the less favourable cardiac remodelling observed in postmenopausal women appears to be driven predominantly by a greater burden of cardiometabolic risk factors, particularly elevated blood pressure, increased adiposity, and metabolic dysfunction, rather than by increased myocardial susceptibility to these exposures.

### Risk factors and CMR according to menopause

In our study, women showed a steady increase in BMI with age and more pronounced midlife increases in systolic blood pressure, LDL-C, and triglycerides, whereas men exhibited earlier but more stable increases, with several measures plateauing in older adulthood. Although these patterns reflect age-related differences observed cross-sectionally, they are consistent with evidence from large longitudinal cohorts describing sex-specific cardiometabolic trajectories across adulthood. In a combined analysis of community-based cohorts, women experienced steeper age-related increases in systolic blood pressure than men across the life course (20), and longitudinal data from the Study of Women’s Health Across the Nation indicate that LDL-C and triglyceride levels rise across the menopausal transition independent of age (21).

Cardiac volumes decreased with age in both sexes, supporting previous findings (15). However, the rate of decline in women became steeper in midlife, whereas in men it remained relatively consistent across the life course. Structural parameters such as LVWT and LVCI increased with age in both groups, with women exhibiting a sharper midlife slope than men. The divergence in midlife trajectories between women and men suggests a role for menopause in accelerating cardiac remodelling in women. This interpretation is supported by findings from the UK Biobank, which show that among postmenopausal women, earlier menopause and a longer postmenopausal interval are associated with smaller LVEDV and more concentric ventricular geometry (22). In addition, longitudinal observations from the Framingham Heart Study reported steeper age-related increases in LVWT in women based on echocardiography (3).

### Metabolic risk factors

In premenopausal women, higher BMI was associated with greater LV mass, larger chamber volumes, and increased stroke volume, with preserved EF and concentricity, suggesting compensatory chamber enlargement (23, 24). In contrast, in postmenopausal women, higher BMI was more strongly associated with increased LVCI and lower LVEF, indicating a shift toward less favourable myocardial remodelling and functional impairment (23, 24). We hypothesize that changes in body composition which accompany menopausal transition such as increasing visceral adiposity and a decline in lean mass (25), modify the association between BMI and myocardial function, such that identical BMI values might reflect a more unfavourable adipose tissue distribution in older women. Visceral adiposity has been implicated in adverse cardiac remodelling through haemodynamic loading and inflammation-mediated pathways that promote myocardial structural change and functional impairment (26), providing a plausible context for these findings.

Previous studies suggested that men predominantly develop eccentric remodelling and women a more concentric response to increased afterload (27), but recent imaging studies have indicated that blood-pressure related increases in concentricity may occur in both sexes (28), though evidence remains limited. In our study, systolic blood pressure showed stronger associations with LV mass and chamber volumes in men, consistent with a more eccentric pattern. However, associations of systolic blood pressure with concentric indices like LVCI and LVWT, were similar between sexes, suggesting that pressure overload promotes concentric adaptation in both women and men, with men exhibiting additional chamber enlargement and mass increases.

Large population-based studies with long-term follow-up have shown that triglyceride levels predict cardiovascular mortality and events in both sexes, with steeper risk gradients in women. In both Norwegian and Danish cohorts followed for nearly three decades, higher triglyceride levels were associated with increased risk of cardiovascular events and mortality, with larger hazard ratios per unit increase in women than in men (29, 30). Consistent with these population-level observations, we found that higher triglyceride levels were associated with lower cardiac volumes and reduced CO in both sexes, together with higher LVCI and LVWT. These associations were consistently strongest in premenopausal women and significantly exceeded those observed in men under 50 years of age, particularly for the right ventricle. As triglyceride concentrations are widely regarded as an integrated marker of metabolic impairment (31), the lower baseline levels in premenopausal women mean that an equivalent absolute increase may reflect proportionally greater disturbance in insulin sensitivity and lipid metabolism, potentially contributing to the stronger associations observed.

### Lifestyle risk factors

Smoking is an impactful cardiovascular lifestyle risk factor, with well-established effects on cardiac remodelling (32). In our cohort, differences in associations of smoking status with CMR attenuated after accounting for cumulative exposure. Younger participants were more likely to be current smokers but had lower lifetime exposure, whereas older participants were more often former smokers with substantially higher cumulative exposure, suggesting that the observed differences were driven more by cumulative smoking burden rather than age-related differences in biological vulnerability. Further studies are needed to determine how smoking intensity, duration, and timing across the life course affect cardiac remodelling.

Physical activity was associated with modest increases in cardiac volumes, outputs, and left ventricular mass, without accompanying changes in EF or concentricity, a pattern consistent with physiological rather than pathological remodelling (33). Men below 50 years demonstrated a noticeably stronger remodelling association than older men and premenopausal women. One possible explanation is that younger men tend to gain more lean muscle mass in response to physical activity than either older men (34) or women (35), and higher lean mass is in turn associated with larger cardiac size, including greater left ventricular mass and chamber dimensions (36).

Our findings with regards to sleep relate to previous large-scale population work which suggests that long sleep duration may have more adverse cardiac effects in men than in women (37). In our study, where we modelled both sleep duration and perceived sleep problems simultaneously, long sleep duration showed broadly similar associations in men and women, while sleep problems demonstrated stronger adverse relationships in men. Overall, the combined evidence suggests that men may be more susceptible to certain adverse sleep-related cardiac changes, potentially due to physiological or hormonal differences. There is evidence which indicates that sleep restriction can disrupt endocrine function, including elevation of cortisol (38) and reductions in testosterone levels (39). Lower testosterone levels have been linked to poorer cardiovascular health and increased cardiovascular risk in men (40), providing a possible pathway through which sleep problems may exert stronger cardiac effects in men. Nonetheless, the mechanisms underlying the sex differences in the contributions of sleep duration and sleep problems need further investigation.

### Strengths and Limitations

A strength of our study is the large, population-based dataset with a sufficiently broad age range to examine cardiac remodelling across different life stages, including premenopausal and postmenopausal women and the male counterparts of matching age. In addition, cardiac phenotyping was performed using cardiac MRI, the gold-standard modality for quantifying ventricular structure and function, allowing precise assessment of both the left and right ventricles. A limitation of our study is that menopausal status could not be defined with high precision, as the available data included only self-reported information and age-based criteria rather than hormonal measurements. This may have introduced some misclassification. Additionally, since the study was cross-sectional, we cannot determine the direction of the observed associations or draw definitive conclusions about causality.

### Conclusion/Outlook

Our study suggests that adverse cardiac remodelling occurs in individuals with a higher burden of cardiovascular risk factors regardless of age. Although both cardiometabolic profiles and cardiac parameters vary substantially with age, the strength of their associations differed more markedly between sexes than between younger and older adults. In the context of menopause, our findings suggest that postmenopausal differences in cardiac remodelling are driven primarily by a higher burden of cardiometabolic risk factors, including blood pressure, adiposity, and metabolic health, rather than by altered cardiac responsiveness to these risk factors. From a public health perspective, these findings underscore the importance of effective sex-specific cardiometabolic risk factor control across the life course.

## Abbreviations

BMI: body mass index
BP: blood pressure
CI: confidence interval
CMR: cardiac magnetic resonance
CO: cardiac output
CVD: cardiovascular disease
EDV: end-diastolic volume
EF: ejection fraction
ESV: end-systolic volume
FDR: false discovery rate
GAM: generalized additive models
HbA1c: haemoglobin A1c
HDL-C: high-density lipoprotein cholesterol
HFpEF: heart failure with preserved ejection fraction
LDL-C: low-density lipoprotein cholesterol
LV: left ventricle
LVCI: left ventricular concentricity index
LVM: left ventricular mass
LVWT: left ventricular wall thickness
MET: metabolic equivalent of task
MRI: magnetic resonance imaging
NAKO: German National Cohort
RV: right ventricle
SD: standard deviation
SV: stroke volume.

## Declarations

### Funding/ Acknowledgements

This project was conducted with data (application no. NAKO-745) from the German National Cohort (NAKO). The NAKO is funded by the Federal Ministry of Education and Research (BMBF) (project funding reference numbers: 01ER1301A/B/C, 01ER1511D, 01ER1801A/B/C/D and 01ER2301A/B/C), federal states of Germany and the Helmholtz Association, the participating universities, and the institutes of the Leibniz Association. This project received funding from the Deutsche Forschungsgemeinschaft (DFG, German Research Foundation) – project no. 428224476 / SPP 2177. We thank all participants who took part in the NAKO study and the staff of this research initiative.

PMF has received the Kaltenbach Scholarship from the German Heart Foundation.

### Data availability

Access to and use of NAKO (German National Cohort) data and biosamples can be obtained via an electronic application portal (https://transfer.nako.de/transfer/index).

### Credit Author statement

Conceptualization: MF, SR; Investigation: MF, CS, PMF, JM, RTS, MR, JSM, CLS, SR; Methodology: MF, SR; Formal analysis: MF; Resources: MF, CS, PMF, JM, RTS, MD, JG, TK, ML, WL, FN, KS, MR, FB, JSM, CLS, SR; Data Curation: MF, CS, RTS, SR; Visualization: MF; Writing - original draft: MF, SR; Writing - review and editing: MF, CS, PMF, JM, RTS, MD, JG, TK, ML, WL, FN, KS, MR, FB, JSM, CLS, SR

