## Supplementary Material for "Sex- and age-related cardiac remodelling and its association with risk factors – Results from Cardiovascular Magnetic Resonance Imaging in the German National Cohort (NAKO)"

Flis et al.

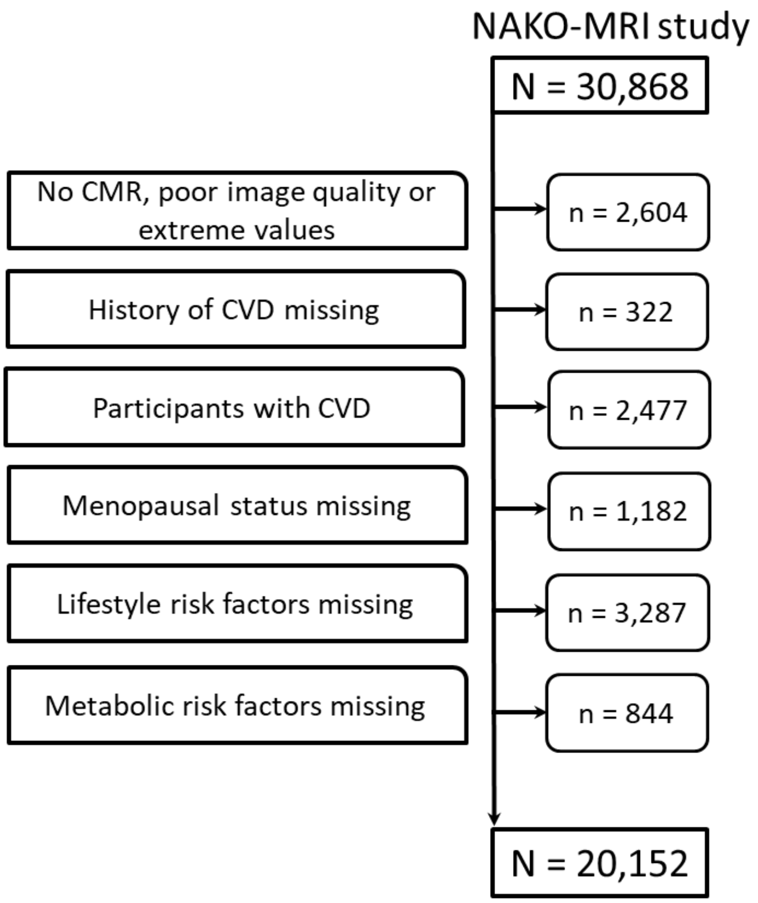


**Supplementary Figure 1.** Participant flowchart. CMR, cardiac magnetic resonance; CVD, cardiovascular disease (defined as self-reported physician diagnosis of myocardial infarction, coronary heart disease, heart failure, arrhythmias, and peripheral arterial disease)

**Supplementary Table 1.** Association of body mass index with cardiac parameters, and FDR-adjusted Wald test p-values for differences in regression coefficients

| | Women | | | | | | Men | | | | | | Wald test | | | |
|---|---|---|---|---|---|---|---|---|---|---|---|---|---|---|---|---|
| | Premenopausal | | | Postmenopausal | | | Age<50 | | | Age≥50 | | | | | | |
| **Outcome** | **Beta** | **95% CI** | **p-value** | **Beta** | **95% CI** | **p-value** | **Beta** | **95% CI** | **p-value** | **Beta** | **95% CI** | **p-value** | **p-value[a]** | **p-value[b]** | **p-value[c]** | **p-value[d]** |
| LV Ejection Fraction | -0.009 | -0.036, 0.018 | 0.505 | -0.090 | -0.123, -0.057 | <0.001 | -0.048 | -0.077, -0.019 | 0.001 | -0.087 | -0.126, -0.048 | <0.001 | 0.005 | 0.316 | 0.206 | 0.946 |
| LV End-diastolic Volume | 0.306 | 0.283, 0.329 | <0.001 | 0.241 | 0.214, 0.268 | <0.001 | 0.274 | 0.245, 0.302 | <0.001 | 0.230 | 0.193, 0.267 | <0.001 | 0.006 | 0.236 | 0.263 | 0.809 |
| LV End-systolic Volume | 0.227 | 0.203, 0.250 | <0.001 | 0.220 | 0.192, 0.247 | <0.001 | 0.243 | 0.214, 0.272 | <0.001 | 0.231 | 0.194, 0.269 | <0.001 | 0.834 | 0.809 | 0.652 | 0.803 |
| LV Stroke Volume | 0.289 | 0.265, 0.314 | <0.001 | 0.198 | 0.170, 0.227 | <0.001 | 0.230 | 0.201, 0.259 | <0.001 | 0.174 | 0.137, 0.211 | <0.001 | <0.001 | 0.109 | 0.030 | 0.566 |
| LV Cardiac Output | 0.215 | 0.188, 0.242 | <0.001 | 0.174 | 0.143, 0.204 | <0.001 | 0.186 | 0.156, 0.216 | <0.001 | 0.135 | 0.097, 0.172 | <0.001 | 0.183 | 0.158 | 0.381 | 0.322 |
| RV Ejection Fraction | -0.064 | -0.093, -0.036 | <0.001 | -0.043 | -0.077, -0.010 | 0.011 | -0.102 | -0.131, -0.074 | <0.001 | -0.079 | -0.115, -0.042 | <0.001 | 0.612 | 0.584 | 0.236 | 0.386 |
| RV End-diastolic Volume | 0.284 | 0.261, 0.307 | <0.001 | 0.207 | 0.182, 0.233 | <0.001 | 0.280 | 0.252, 0.308 | <0.001 | 0.232 | 0.197, 0.267 | <0.001 | <0.001 | 0.155 | 0.921 | 0.522 |
| RV End-systolic Volume | 0.235 | 0.211, 0.259 | <0.001 | 0.162 | 0.137, 0.186 | <0.001 | 0.279 | 0.249, 0.308 | <0.001 | 0.216 | 0.182, 0.250 | <0.001 | <0.001 | 0.056 | 0.123 | 0.075 |
| RV Stroke Volume | 0.225 | 0.200, 0.251 | <0.001 | 0.174 | 0.144, 0.205 | <0.001 | 0.177 | 0.148, 0.207 | <0.001 | 0.161 | 0.123, 0.199 | <0.001 | 0.082 | 0.743 | 0.100 | 0.785 |
| RV Cardiac Output | 0.170 | 0.142, 0.197 | <0.001 | 0.156 | 0.125, 0.187 | <0.001 | 0.146 | 0.116, 0.177 | <0.001 | 0.126 | 0.088, 0.165 | <0.001 | 0.751 | 0.681 | 0.515 | 0.500 |
| LV Concentricity Index | 0.047 | 0.031, 0.063 | <0.001 | 0.097 | 0.072, 0.121 | <0.001 | 0.150 | 0.127, 0.174 | <0.001 | 0.176 | 0.139, 0.213 | <0.001 | 0.016 | 0.501 | <0.001 | 0.008 |
| LV Wall Thickness | 0.272 | 0.254, 0.290 | <0.001 | 0.310 | 0.284, 0.336 | <0.001 | 0.346 | 0.324, 0.368 | <0.001 | 0.391 | 0.359, 0.424 | <0.001 | 0.109 | 0.123 | <0.001 | 0.003 |
| LV Mass | 0.323 | 0.308, 0.338 | <0.001 | 0.322 | 0.303, 0.341 | <0.001 | 0.444 | 0.423, 0.465 | <0.001 | 0.445 | 0.417, 0.473 | <0.001 | 0.957 | 0.988 | <0.001 | <0.001 |

Results from fully adjusted models. β coefficients are reported per 1-SD increase in continuous exposures, with outcomes standardized. P-values were obtained from Wald tests and adjusted using the false discovery rate (FDR). [a]Premenopausal women vs Postmenopausal women; [b]Men < 50 years vs Men ≥ 50 years; [c]Premenopausal Women vs Men < 50 years; [d]Postmenopausal women vs Men ≥ 50 years. CI, confidence interval; LV, left ventricle; RV, right ventricle

**Supplementary Table 2.** Association of systolic blood pressure with cardiac parameters, and FDR-adjusted Wald test p-values for differences in regression coefficients

| | Women | | | | | | Men | | | | | | Wald test | | | |
|---|---|---|---|---|---|---|---|---|---|---|---|---|---|---|---|---|
| | Premenopausal | | | Postmenopausal | | | Age<50 | | | Age≥50 | | | | | | |
| **Outcome** | **Beta** | **95% CI** | **p-value** | **Beta** | **95% CI** | **p-value** | **Beta** | **95% CI** | **p-value** | **Beta** | **95% CI** | **p-value** | **p-value[a]** | **p-value[b]** | **p-value[c]** | **p-value[d]** |
| LV Ejection Fraction | 0.070 | 0.019, 0.120 | 0.007 | 0.100 | 0.055, 0.144 | <0.001 | 0.161 | 0.121, 0.202 | <0.001 | 0.165 | 0.124, 0.206 | <0.001 | 0.630 | 0.946 | 0.052 | 0.152 |
| LV End-diastolic Volume | 0.143 | 0.100, 0.185 | <0.001 | 0.140 | 0.104, 0.176 | <0.001 | 0.231 | 0.190, 0.271 | <0.001 | 0.207 | 0.168, 0.246 | <0.001 | 0.950 | 0.662 | 0.037 | 0.088 |
| LV End-systolic Volume | 0.066 | 0.023, 0.109 | 0.003 | 0.052 | 0.016, 0.089 | 0.005 | 0.082 | 0.041, 0.124 | <0.001 | 0.066 | 0.026, 0.105 | 0.001 | 0.809 | 0.782 | 0.793 | 0.806 |
| LV Stroke Volume | 0.164 | 0.118, 0.209 | <0.001 | 0.169 | 0.131, 0.207 | <0.001 | 0.282 | 0.241, 0.324 | <0.001 | 0.259 | 0.220, 0.299 | <0.001 | 0.926 | 0.681 | 0.003 | 0.020 |
| LV Cardiac Output | 0.119 | 0.068, 0.169 | <0.001 | 0.124 | 0.083, 0.165 | <0.001 | 0.218 | 0.176, 0.261 | <0.001 | 0.202 | 0.162, 0.241 | <0.001 | 0.939 | 0.783 | 0.036 | 0.063 |
| RV Ejection Fraction | 0.173 | 0.120, 0.227 | <0.001 | 0.134 | 0.090, 0.179 | <0.001 | 0.156 | 0.116, 0.197 | <0.001 | 0.175 | 0.137, 0.214 | <0.001 | 0.517 | 0.746 | 0.803 | 0.399 |
| RV End-diastolic Volume | 0.089 | 0.047, 0.132 | <0.001 | 0.065 | 0.031, 0.100 | <0.001 | 0.174 | 0.135, 0.214 | <0.001 | 0.113 | 0.076, 0.151 | <0.001 | 0.640 | 0.140 | 0.046 | 0.230 |
| RV End-systolic Volume | -0.034 | -0.079, 0.010 | 0.130 | -0.025 | -0.057, 0.007 | 0.127 | 0.039 | -0.002, 0.080 | 0.066 | -0.022 | -0.058, 0.014 | 0.231 | 0.858 | 0.140 | 0.108 | 0.944 |
| RV Stroke Volume | 0.175 | 0.128, 0.223 | <0.001 | 0.128 | 0.088, 0.168 | <0.001 | 0.240 | 0.199, 0.282 | <0.001 | 0.201 | 0.161, 0.242 | <0.001 | 0.357 | 0.428 | 0.176 | 0.082 |
| RV Cardiac Output | 0.130 | 0.079, 0.180 | <0.001 | 0.091 | 0.050, 0.133 | <0.001 | 0.191 | 0.148, 0.234 | <0.001 | 0.158 | 0.117, 0.199 | <0.001 | 0.505 | 0.544 | 0.240 | 0.124 |
| LV Concentricity Index | 0.105 | 0.075, 0.135 | <0.001 | 0.040 | 0.007, 0.072 | 0.017 | 0.097 | 0.064, 0.130 | <0.001 | 0.072 | 0.032, 0.111 | <0.001 | 0.040 | 0.602 | 0.845 | 0.457 |
| LV Wall Thickness | 0.164 | 0.129, 0.198 | <0.001 | 0.142 | 0.108, 0.177 | <0.001 | 0.183 | 0.152, 0.215 | <0.001 | 0.194 | 0.160, 0.229 | <0.001 | 0.640 | 0.809 | 0.666 | 0.155 |
| LV Mass | 0.229 | 0.201, 0.258 | <0.001 | 0.175 | 0.149, 0.201 | <0.001 | 0.351 | 0.322, 0.381 | <0.001 | 0.301 | 0.271, 0.331 | <0.001 | 0.051 | 0.108 | <0.001 | <0.001 |

Results from fully adjusted models. β coefficients are reported per 1-SD increase in continuous exposures, with outcomes standardized. P-values were obtained from Wald tests and adjusted using the false discovery rate (FDR). [a]Premenopausal women vs Postmenopausal women; [b]Men < 50 years vs Men ≥ 50 years; [c]Premenopausal Women vs Men < 50 years; [d]Postmenopausal women vs Men ≥ 50 years. CI, confidence interval; LV, left ventricle; RV, right ventricle

**Supplementary Table 3.** Association of diastolic blood pressure with cardiac parameters, and FDR-adjusted Wald test p-values for differences in regression coefficients

| | Women | | | | | | Men | | | | | | Wald test | | | |
|---|---|---|---|---|---|---|---|---|---|---|---|---|---|---|---|---|
| | Premenopausal | | | Postmenopausal | | | Age<50 | | | Age≥50 | | | | | | |
| **Outcome** | **Beta** | **95% CI** | **p-value** | **Beta** | **95% CI** | **p-value** | **Beta** | **95% CI** | **p-value** | **Beta** | **95% CI** | **p-value** | **p-value[a]** | **p-value[b]** | **p-value[c]** | **p-value[d]** |
| LV Ejection Fraction | -0.053 | -0.098, -0.007 | 0.023 | -0.103 | -0.152, -0.054 | <0.001 | -0.126 | -0.162, -0.089 | <0.001 | -0.194 | -0.236, -0.151 | <0.001 | 0.359 | 0.105 | 0.090 | 0.058 |
| LV End-diastolic Volume | -0.181 | -0.219, -0.143 | <0.001 | -0.159 | -0.199, -0.119 | <0.001 | -0.316 | -0.353, -0.280 | <0.001 | -0.237 | -0.277, -0.197 | <0.001 | 0.686 | 0.043 | <0.001 | 0.061 |
| LV End-systolic Volume | -0.102 | -0.141, -0.063 | <0.001 | -0.061 | -0.102, -0.021 | 0.003 | -0.169 | -0.206, -0.131 | <0.001 | -0.071 | -0.112, -0.030 | <0.001 | 0.385 | 0.011 | 0.097 | 0.853 |
| LV Stroke Volume | -0.195 | -0.235, -0.154 | <0.001 | -0.191 | -0.233, -0.149 | <0.001 | -0.347 | -0.384, -0.310 | <0.001 | -0.299 | -0.340, -0.259 | <0.001 | 0.947 | 0.273 | <0.001 | 0.006 |
| LV Cardiac Output | -0.112 | -0.157, -0.066 | <0.001 | -0.108 | -0.153, -0.062 | <0.001 | -0.249 | -0.287, -0.211 | <0.001 | -0.236 | -0.277, -0.195 | <0.001 | 0.946 | 0.809 | <0.001 | <0.001 |
| RV Ejection Fraction | -0.053 | -0.101, -0.005 | 0.029 | -0.039 | -0.088, 0.011 | 0.128 | -0.069 | -0.105, -0.032 | <0.001 | -0.113 | -0.153, -0.074 | <0.001 | 0.820 | 0.301 | 0.803 | 0.116 |
| RV End-diastolic Volume | -0.180 | -0.218, -0.141 | <0.001 | -0.119 | -0.157, -0.081 | <0.001 | -0.271 | -0.307, -0.236 | <0.001 | -0.212 | -0.251, -0.173 | <0.001 | 0.140 | 0.135 | 0.011 | 0.014 |
| RV End-systolic Volume | -0.097 | -0.137, -0.057 | <0.001 | -0.063 | -0.099, -0.027 | <0.001 | -0.168 | -0.205, -0.131 | <0.001 | -0.089 | -0.126, -0.052 | <0.001 | 0.455 | 0.036 | 0.077 | 0.585 |
| RV Stroke Volume | -0.193 | -0.236, -0.150 | <0.001 | -0.129 | -0.173, -0.084 | <0.001 | -0.270 | -0.307, -0.233 | <0.001 | -0.252 | -0.293, -0.210 | <0.001 | 0.173 | 0.751 | 0.063 | 0.003 |
| RV Cardiac Output | -0.117 | -0.162, -0.071 | <0.001 | -0.058 | -0.104, -0.012 | 0.014 | -0.196 | -0.234, -0.158 | <0.001 | -0.201 | -0.244, -0.159 | <0.001 | 0.249 | 0.926 | 0.073 | <0.001 |
| LV Concentricity Index | 0.092 | 0.065, 0.119 | <0.001 | 0.114 | 0.077, 0.150 | <0.001 | 0.153 | 0.124, 0.183 | <0.001 | 0.160 | 0.119, 0.201 | <0.001 | 0.613 | 0.896 | 0.036 | 0.283 |
| LV Wall Thickness | 0.057 | 0.026, 0.087 | <0.001 | 0.068 | 0.030, 0.106 | <0.001 | 0.063 | 0.035, 0.091 | <0.001 | 0.053 | 0.018, 0.089 | 0.003 | 0.810 | 0.821 | 0.875 | 0.785 |
| LV Mass | -0.062 | -0.087, -0.036 | <0.001 | -0.044 | -0.072, -0.015 | 0.003 | -0.155 | -0.182, -0.129 | <0.001 | -0.089 | -0.120, -0.058 | <0.001 | 0.613 | 0.022 | <0.001 | 0.148 |

Results from fully adjusted models. β coefficients are reported per 1-SD increase in continuous exposures, with outcomes standardized. P-values were obtained from Wald tests and adjusted using the false discovery rate (FDR). [a]Premenopausal women vs Postmenopausal women; [b]Men < 50 years vs Men ≥ 50 years; [c]Premenopausal Women vs Men < 50 years; [d]Postmenopausal women vs Men ≥ 50 years. CI, confidence interval; LV, left ventricle; RV, right ventricle

**Supplementary Table 4.** Association of low-density lipoprotein with cardiac parameters, and FDR-adjusted Wald test p-values for differences in regression coefficients

| | Women | | | | | | Men | | | | | | Wald test | | | |
|---|---|---|---|---|---|---|---|---|---|---|---|---|---|---|---|---|
| | Premenopausal | | | Postmenopausal | | | Age<50 | | | Age≥50 | | | | | | |
| **Outcome** | **Beta** | **95% CI** | **p-value** | **Beta** | **95% CI** | **p-value** | **Beta** | **95% CI** | **p-value** | **Beta** | **95% CI** | **p-value** | **p-value[a]** | **p-value[b]** | **p-value[c]** | **p-value[d]** |
| LV Ejection Fraction | -0.008 | -0.041, 0.025 | 0.638 | -0.004 | -0.039, 0.030 | 0.803 | 0.012 | -0.013, 0.038 | 0.339 | -0.004 | -0.036, 0.028 | 0.818 | 0.939 | 0.686 | 0.600 | 0.991 |
| LV End-diastolic Volume | -0.050 | -0.077, -0.022 | <0.001 | -0.044 | -0.072, -0.016 | 0.002 | -0.096 | -0.121, -0.070 | <0.001 | -0.075 | -0.105, -0.045 | <0.001 | 0.888 | 0.573 | 0.101 | 0.359 |
| LV End-systolic Volume | -0.035 | -0.063, -0.007 | 0.015 | -0.028 | -0.057, 0.000 | 0.054 | -0.081 | -0.107, -0.055 | <0.001 | -0.052 | -0.083, -0.022 | <0.001 | 0.857 | 0.388 | 0.112 | 0.515 |
| LV Stroke Volume | -0.048 | -0.078, -0.019 | 0.001 | -0.045 | -0.075, -0.015 | 0.003 | -0.083 | -0.109, -0.057 | <0.001 | -0.074 | -0.104, -0.044 | <0.001 | 0.939 | 0.809 | 0.255 | 0.418 |
| LV Cardiac Output | -0.023 | -0.057, 0.010 | 0.166 | -0.047 | -0.079, -0.014 | 0.005 | -0.050 | -0.077, -0.024 | <0.001 | -0.047 | -0.077, -0.016 | 0.003 | 0.590 | 0.928 | 0.456 | 0.998 |
| RV Ejection Fraction | 0.011 | -0.024, 0.046 | 0.533 | -0.020 | -0.054, 0.015 | 0.274 | 0.009 | -0.017, 0.034 | 0.512 | -0.021 | -0.051, 0.009 | 0.167 | 0.466 | 0.359 | 0.948 | 0.971 |
| RV End-diastolic Volume | -0.025 | -0.053, 0.003 | 0.078 | -0.053 | -0.080, -0.026 | <0.001 | -0.067 | -0.092, -0.042 | <0.001 | -0.050 | -0.079, -0.021 | <0.001 | 0.381 | 0.630 | 0.139 | 0.936 |
| RV End-systolic Volume | -0.026 | -0.055, 0.003 | 0.083 | -0.030 | -0.055, -0.004 | 0.022 | -0.057 | -0.082, -0.031 | <0.001 | -0.023 | -0.050, 0.005 | 0.107 | 0.921 | 0.255 | 0.322 | 0.845 |
| RV Stroke Volume | -0.015 | -0.046, 0.016 | 0.330 | -0.056 | -0.088, -0.025 | <0.001 | -0.053 | -0.079, -0.026 | <0.001 | -0.058 | -0.089, -0.027 | <0.001 | 0.237 | 0.899 | 0.244 | 0.971 |
| RV Cardiac Output | 0.000 | -0.033, 0.033 | 0.983 | -0.056 | -0.088, -0.023 | <0.001 | -0.029 | -0.057, -0.002 | 0.033 | -0.036 | -0.067, -0.004 | 0.028 | 0.109 | 0.881 | 0.417 | 0.632 |
| LV Concentricity Index | 0.012 | -0.008, 0.031 | 0.244 | 0.027 | 0.001, 0.052 | 0.040 | 0.026 | 0.005, 0.047 | 0.017 | 0.016 | -0.014, 0.046 | 0.301 | 0.613 | 0.803 | 0.602 | 0.793 |
| LV Wall Thickness | 0.009 | -0.013, 0.031 | 0.426 | 0.010 | -0.017, 0.037 | 0.484 | 0.001 | -0.019, 0.020 | 0.951 | -0.015 | -0.041, 0.012 | 0.278 | 0.990 | 0.619 | 0.785 | 0.443 |
| LV Mass | -0.034 | -0.052, -0.015 | <0.001 | -0.017 | -0.037, 0.004 | 0.106 | -0.071 | -0.090, -0.052 | <0.001 | -0.064 | -0.087, -0.041 | <0.001 | 0.466 | 0.809 | 0.052 | 0.033 |

Results from fully adjusted models. β coefficients are reported per 1-SD increase in continuous exposures, with outcomes standardized. P-values were obtained from Wald tests and adjusted using the false discovery rate (FDR). [a]Premenopausal women vs Postmenopausal women; [b]Men < 50 years vs Men ≥ 50 years; [c]Premenopausal Women vs Men < 50 years; [d]Postmenopausal women vs Men ≥ 50 years. CI, confidence interval; LV, left ventricle; RV, right ventricle

**Supplementary Table 5.** Association of triglycerides with cardiac parameters, and FDR-adjusted Wald test p-values for differences in regression coefficients

| | Women | | | | | | Men | | | | | | Wald test | | | |
|---|---|---|---|---|---|---|---|---|---|---|---|---|---|---|---|---|
| | Premenopausal | | | Postmenopausal | | | Age<50 | | | Age≥50 | | | | | | |
| **Outcome** | **Beta** | **95% CI** | **p-value** | **Beta** | **95% CI** | **p-value** | **Beta** | **95% CI** | **p-value** | **Beta** | **95% CI** | **p-value** | **p-value[a]** | **p-value[b]** | **p-value[c]** | **p-value[d]** |
| LV Ejection Fraction | 0.008 | -0.039, 0.055 | 0.739 | 0.047 | -0.005, 0.098 | 0.076 | -0.003 | -0.028, 0.022 | 0.813 | 0.015 | -0.015, 0.046 | 0.320 | 0.530 | 0.613 | 0.829 | 0.567 |
| LV End-diastolic Volume | -0.171 | -0.211, -0.131 | <0.001 | -0.120 | -0.162, -0.078 | <0.001 | -0.086 | -0.111, -0.061 | <0.001 | -0.071 | -0.100, -0.042 | <0.001 | 0.261 | 0.697 | 0.008 | 0.226 |
| LV End-systolic Volume | -0.127 | -0.168, -0.086 | <0.001 | -0.111 | -0.154, -0.069 | <0.001 | -0.063 | -0.089, -0.038 | <0.001 | -0.062 | -0.091, -0.033 | <0.001 | 0.798 | 0.971 | 0.075 | 0.226 |
| LV Stroke Volume | -0.162 | -0.204, -0.119 | <0.001 | -0.097 | -0.141, -0.053 | <0.001 | -0.082 | -0.107, -0.056 | <0.001 | -0.061 | -0.090, -0.032 | <0.001 | 0.164 | 0.546 | 0.023 | 0.414 |
| LV Cardiac Output | -0.106 | -0.154, -0.058 | <0.001 | -0.092 | -0.140, -0.043 | <0.001 | -0.048 | -0.074, -0.022 | <0.001 | -0.020 | -0.050, 0.009 | 0.174 | 0.825 | 0.405 | 0.158 | 0.090 |
| RV Ejection Fraction | 0.054 | 0.004, 0.104 | 0.035 | 0.063 | 0.011, 0.115 | 0.018 | 0.001 | -0.024, 0.026 | 0.937 | 0.008 | -0.021, 0.036 | 0.599 | 0.901 | 0.852 | 0.230 | 0.236 |
| RV End-diastolic Volume | -0.173 | -0.213, -0.132 | <0.001 | -0.112 | -0.152, -0.072 | <0.001 | -0.064 | -0.088, -0.039 | <0.001 | -0.050 | -0.078, -0.022 | <0.001 | 0.155 | 0.707 | <0.001 | 0.090 |
| RV End-systolic Volume | -0.153 | -0.195, -0.112 | <0.001 | -0.106 | -0.144, -0.068 | <0.001 | -0.050 | -0.075, -0.024 | <0.001 | -0.042 | -0.069, -0.016 | 0.002 | 0.288 | 0.830 | <0.001 | 0.060 |
| RV Stroke Volume | -0.127 | -0.172, -0.082 | <0.001 | -0.076 | -0.123, -0.029 | 0.002 | -0.054 | -0.080, -0.028 | <0.001 | -0.039 | -0.069, -0.009 | 0.010 | 0.324 | 0.707 | 0.052 | 0.434 |
| RV Cardiac Output | -0.080 | -0.128, -0.033 | <0.001 | -0.078 | -0.126, -0.029 | 0.002 | -0.029 | -0.056, -0.003 | 0.031 | -0.007 | -0.038, 0.023 | 0.633 | 0.968 | 0.544 | 0.233 | 0.100 |
| LV Concentricity Index | 0.084 | 0.056, 0.112 | <0.001 | 0.076 | 0.038, 0.114 | <0.001 | 0.052 | 0.031, 0.073 | <0.001 | 0.071 | 0.041, 0.100 | <0.001 | 0.857 | 0.572 | 0.244 | 0.91 |
| LV Wall Thickness | 0.034 | 0.002, 0.066 | 0.038 | 0.009 | -0.031, 0.049 | 0.655 | 0.025 | 0.006, 0.045 | 0.011 | 0.040 | 0.015, 0.066 | 0.002 | 0.602 | 0.613 | 0.809 | 0.434 |
| LV Mass | -0.065 | -0.091, -0.038 | <0.001 | -0.052 | -0.082, -0.022 | <0.001 | -0.037 | -0.055, -0.019 | <0.001 | -0.011 | -0.033, 0.011 | 0.340 | 0.758 | 0.241 | 0.285 | 0.146 |

Results from fully adjusted models. β coefficients are reported per 1-SD increase in continuous exposures, with outcomes standardized. P-values were obtained from Wald tests and adjusted using the false discovery rate (FDR). [a]Premenopausal women vs Postmenopausal women; [b]Men < 50 years vs Men ≥ 50 years; [c]Premenopausal Women vs Men < 50 years; [d]Postmenopausal women vs Men ≥ 50 years. CI, confidence interval; LV, left ventricle; RV, right ventricle

**Supplementary Table 6.** Association of haemoglobin A1c with cardiac parameters, and FDR-adjusted Wald test p-values for differences in regression coefficients

| | Women | | | | | | Men | | | | | | Wald test (p-value) | | | |
|---|---|---|---|---|---|---|---|---|---|---|---|---|---|---|---|---|
| | Premenopausal | | | Postmenopausal | | | Age<50 | | | Age≥50 | | | | | | |
| **Outcome** | **Beta** | **95% CI** | **p-value** | **Beta** | **95% CI** | **p-value** | **Beta** | **95% CI** | **p-value** | **Beta** | **95% CI** | **p-value** | **p-value[a]** | **p-value[b]** | **p-value[c]** | **p-value[d]** |
| LV Ejection Fraction | -0.019 | -0.056, 0.018 | 0.315 | -0.008 | -0.051, 0.035 | 0.718 | -0.047 | -0.078, -0.016 | 0.003 | -0.026 | -0.057, 0.006 | 0.107 | 0.834 | 0.613 | 0.512 | 0.746 |
| LV End-diastolic Volume | -0.019 | -0.050, 0.012 | 0.237 | -0.034 | -0.069, 0.001 | 0.057 | -0.041 | -0.072, -0.010 | 0.009 | -0.063 | -0.093, -0.033 | <0.001 | 0.759 | 0.572 | 0.585 | 0.443 |
| LV End-systolic Volume | -0.002 | -0.034, 0.030 | 0.924 | -0.025 | -0.061, 0.010 | 0.162 | -0.004 | -0.035, 0.027 | 0.804 | -0.036 | -0.066, -0.005 | 0.021 | 0.593 | 0.381 | 0.949 | 0.814 |
| LV Stroke Volume | -0.027 | -0.060, 0.007 | 0.117 | -0.032 | -0.068, 0.005 | 0.088 | -0.058 | -0.089, -0.027 | <0.001 | -0.068 | -0.098, -0.038 | <0.001 | 0.921 | 0.809 | 0.421 | 0.353 |
| LV Cardiac Output | -0.030 | -0.068, 0.007 | 0.116 | -0.014 | -0.054, 0.025 | 0.476 | -0.028 | -0.060, 0.005 | 0.092 | -0.048 | -0.078, -0.018 | 0.002 | 0.783 | 0.619 | 0.954 | 0.425 |
| RV Ejection Fraction | -0.071 | -0.110, -0.032 | <0.001 | -0.071 | -0.114, -0.028 | 0.001 | -0.013 | -0.044, 0.018 | 0.399 | -0.018 | -0.047, 0.012 | 0.237 | 0.998 | 0.919 | 0.123 | 0.179 |
| RV End-diastolic Volume | -0.007 | -0.038, 0.025 | 0.684 | -0.015 | -0.048, 0.018 | 0.377 | -0.036 | -0.066, -0.006 | 0.019 | -0.052 | -0.080, -0.023 | <0.001 | 0.845 | 0.708 | 0.422 | 0.295 |
| RV End-systolic Volume | 0.038 | 0.005, 0.070 | 0.024 | 0.023 | -0.008, 0.055 | 0.142 | -0.018 | -0.049, 0.013 | 0.249 | -0.030 | -0.057, -0.003 | 0.032 | 0.764 | 0.785 | 0.098 | 0.082 |
| RV Stroke Volume | -0.047 | -0.082, -0.012 | 0.009 | -0.046 | -0.085, -0.008 | 0.019 | -0.040 | -0.072, -0.008 | 0.013 | -0.053 | -0.084, -0.022 | <0.001 | 0.996 | 0.780 | 0.881 | 0.888 |
| RV Cardiac Output | -0.049 | -0.087, -0.012 | 0.009 | -0.024 | -0.064, 0.016 | 0.236 | -0.017 | -0.050, 0.016 | 0.306 | -0.034 | -0.065, -0.003 | 0.034 | 0.619 | 0.707 | 0.434 | 0.839 |
| LV Concentricity Index | 0.025 | 0.003, 0.047 | 0.029 | 0.068 | 0.037, 0.100 | <0.001 | 0.044 | 0.019, 0.070 | <0.001 | 0.056 | 0.026, 0.086 | <0.001 | 0.134 | 0.781 | 0.505 | 0.785 |
| LV Wall Thickness | 0.025 | 0.000, 0.051 | 0.049 | 0.060 | 0.027, 0.093 | <0.001 | 0.041 | 0.017, 0.065 | <0.001 | 0.032 | 0.006, 0.058 | 0.017 | 0.302 | 0.803 | 0.632 | 0.434 |
| LV Mass | 0.010 | -0.011, 0.031 | 0.348 | 0.028 | 0.003, 0.053 | 0.027 | 0.000 | -0.022, 0.022 | 0.993 | -0.019 | -0.041, 0.004 | 0.111 | 0.527 | 0.510 | 0.751 | 0.060 |

Results from fully adjusted models. β coefficients are reported per 1-SD increase in continuous exposures, with outcomes standardized. P-values were obtained from Wald tests and adjusted using the false discovery rate (FDR). [a]Premenopausal women vs Postmenopausal women; [b]Men < 50 years vs Men ≥ 50 years; [c]Premenopausal Women vs Men < 50 years; [d]Postmenopausal women vs Men ≥ 50 years. CI, confidence interval; LV, left ventricle; RV, right ventricle

**Supplementary Table 7.** Association of smoking status: current (compared to never) with cardiac parameters, and FDR-adjusted Wald test p-values for differences in regression coefficients

| | Women | | | | | | Men | | | | | | Wald test | | | |
|---|---|---|---|---|---|---|---|---|---|---|---|---|---|---|---|---|
| | Premenopausal | | | Postmenopausal | | | Age<50 | | | Age≥50 | | | | | | |
| **Outcome** | **Beta** | **95% CI** | **p-value** | **Beta** | **95% CI** | **p-value** | **Beta** | **95% CI** | **p-value** | **Beta** | **95% CI** | **p-value** | **p-value[a]** | **p-value[b]** | **p-value[c]** | **p-value[d]** |
| LV Ejection Fraction | -0.104 | -0.175, -0.032 | 0.004 | -0.068 | -0.161, 0.024 | 0.146 | -0.103 | -0.162, -0.043 | <0.001 | -0.157 | -0.244, -0.070 | <0.001 | 0.780 | 0.571 | 0.992 | 0.399 |
| LV End-diastolic Volume | 0.034 | -0.026, 0.095 | 0.262 | -0.101 | -0.176, -0.026 | 0.008 | -0.084 | -0.144, -0.025 | 0.005 | -0.110 | -0.191, -0.028 | 0.009 | 0.053 | 0.803 | 0.055 | 0.939 |
| LV End-systolic Volume | 0.089 | 0.027, 0.150 | 0.005 | -0.033 | -0.109, 0.043 | 0.396 | 0.007 | -0.054, 0.067 | 0.829 | 0.012 | -0.071, 0.096 | 0.768 | 0.097 | 0.948 | 0.228 | 0.681 |
| LV Stroke Volume | -0.013 | -0.077, 0.052 | 0.697 | -0.126 | -0.205, -0.047 | 0.002 | -0.13 | -0.190, -0.069 | <0.001 | -0.171 | -0.253, -0.089 | <0.001 | 0.140 | 0.681 | 0.074 | 0.687 |
| LV Cardiac Output | -0.027 | -0.099, 0.045 | 0.459 | -0.101 | -0.187, -0.016 | 0.021 | -0.084 | -0.146, -0.021 | 0.008 | -0.110 | -0.194, -0.027 | 0.010 | 0.433 | 0.803 | 0.501 | 0.939 |
| RV Ejection Fraction | 0.002 | -0.074, 0.078 | 0.959 | 0.009 | -0.084, 0.102 | 0.845 | 0.013 | -0.047, 0.073 | 0.670 | -0.014 | -0.095, 0.067 | 0.731 | 0.947 | 0.793 | 0.910 | 0.839 |
| RV End-diastolic Volume | 0.003 | -0.058, 0.064 | 0.916 | -0.120 | -0.191, -0.048 | 0.001 | -0.141 | -0.199, -0.083 | <0.001 | -0.238 | -0.316, -0.159 | <0.001 | 0.077 | 0.198 | 0.014 | 0.140 |
| RV End-systolic Volume | -0.005 | -0.068, 0.058 | 0.881 | -0.089 | -0.156, -0.022 | 0.010 | -0.106 | -0.166, -0.046 | <0.001 | -0.174 | -0.249, -0.099 | <0.001 | 0.244 | 0.388 | 0.123 | 0.288 |
| RV Stroke Volume | 0.010 | -0.058, 0.077 | 0.775 | -0.105 | -0.189, -0.021 | 0.014 | -0.123 | -0.184, -0.062 | <0.001 | -0.211 | -0.296, -0.126 | <0.001 | 0.156 | 0.288 | 0.044 | 0.261 |
| RV Cardiac Output | 0.005 | -0.067, 0.077 | 0.891 | -0.077 | -0.164, 0.009 | 0.080 | -0.077 | -0.140, -0.014 | 0.016 | -0.148 | -0.234, -0.062 | <0.001 | 0.377 | 0.428 | 0.276 | 0.508 |
| LV Concentricity Index | 0.166 | 0.124, 0.209 | <0.001 | 0.342 | 0.274, 0.409 | <0.001 | 0.252 | 0.204, 0.301 | <0.001 | 0.325 | 0.243, 0.408 | <0.001 | <0.001 | 0.353 | 0.074 | 0.876 |
| LV Wall Thickness | 0.224 | 0.175, 0.272 | <0.001 | 0.341 | 0.269, 0.412 | <0.001 | 0.249 | 0.203, 0.295 | <0.001 | 0.317 | 0.246, 0.389 | <0.001 | 0.067 | 0.322 | 0.700 | 0.810 |
| LV Mass | 0.187 | 0.147, 0.227 | <0.001 | 0.206 | 0.153, 0.260 | <0.001 | 0.170 | 0.127, 0.213 | <0.001 | 0.188 | 0.125, 0.250 | <0.001 | 0.782 | 0.809 | 0.782 | 0.810 |

Results from fully adjusted models. β coefficients are reported per 1-SD increase in continuous exposures, with outcomes standardized. P-values were obtained from Wald tests and adjusted using the false discovery rate (FDR). [a]Premenopausal women vs Postmenopausal women; [b]Men < 50 years vs Men ≥ 50 years; [c]Premenopausal Women vs Men < 50 years; [d]Postmenopausal women vs Men ≥ 50 years. CI, confidence interval; LV, left ventricle; RV, right ventricle

**Supplementary Table 8.** Association of smoking status: former (compared to never) with cardiac parameters, and FDR-adjusted Wald test p-values for differences in regression coefficients

| | Women | | | | | | Men | | | | | | Wald test | | | |
|---|---|---|---|---|---|---|---|---|---|---|---|---|---|---|---|---|
| | Premenopausal | | | Postmenopausal | | | Age<50 | | | Age≥50 | | | | | | |
| Outcome | Beta | 95% CI | p-value | Beta | 95% CI | p-value | Beta | 95% CI | p-value | Beta | 95% CI | p-value | p-value[a] | p-value[b] | p-value[c] | p-value[d] |
| LV Ejection Fraction | -0.007 | -0.070, 0.056 | 0.831 | -0.013 | -0.086, 0.059 | 0.718 | 0.015 | -0.040, 0.070 | 0.590 | -0.086 | -0.150, -0.021 | 0.009 | 0.943 | 0.109 | 0.798 | 0.366 |
| LV End-diastolic Volume | 0.049 | -0.004, 0.102 | 0.070 | -0.032 | -0.091, 0.027 | 0.290 | -0.093 | -0.148, -0.039 | <0.001 | -0.066 | -0.127, -0.006 | 0.032 | 0.181 | 0.751 | 0.005 | 0.681 |
| LV End-systolic Volume | 0.041 | -0.013, 0.096 | 0.134 | -0.018 | -0.078, 0.042 | 0.566 | -0.078 | -0.133, -0.022 | 0.006 | 0.000 | -0.062, 0.061 | 0.994 | 0.378 | 0.236 | 0.033 | 0.832 |
| LV Stroke Volume | 0.042 | -0.014, 0.099 | 0.141 | -0.034 | -0.097, 0.028 | 0.276 | -0.082 | -0.137, -0.026 | 0.004 | -0.098 | -0.159, -0.037 | 0.002 | 0.244 | 0.834 | 0.028 | 0.378 |
| LV Cardiac Output | 0.061 | -0.002, 0.124 | 0.059 | -0.023 | -0.090, 0.044 | 0.503 | -0.039 | -0.096, 0.018 | 0.178 | -0.059 | -0.121, 0.002 | 0.060 | 0.246 | 0.809 | 0.116 | 0.686 |
| RV Ejection Fraction | -0.027 | -0.094, 0.039 | 0.423 | -0.044 | -0.117, 0.029 | 0.239 | 0.101 | 0.046, 0.156 | <0.001 | -0.020 | -0.081, 0.040 | 0.505 | 0.857 | 0.038 | 0.038 | 0.803 |
| RV End-diastolic Volume | 0.003 | -0.051, 0.056 | 0.923 | -0.051 | -0.108, 0.005 | 0.074 | -0.100 | -0.153, -0.047 | <0.001 | -0.087 | -0.145, -0.029 | 0.003 | 0.404 | 0.858 | 0.065 | 0.640 |
| RV End-systolic Volume | 0.012 | -0.043, 0.068 | 0.664 | -0.009 | -0.062, 0.044 | 0.741 | -0.141 | -0.196, -0.085 | <0.001 | -0.06 | -0.116, -0.005 | 0.034 | 0.788 | 0.176 | 0.003 | 0.429 |
| RV Stroke Volume | -0.008 | -0.067, 0.052 | 0.801 | -0.073 | -0.139, -0.007 | 0.029 | -0.024 | -0.080, 0.032 | 0.404 | -0.080 | -0.143, -0.018 | 0.012 | 0.370 | 0.424 | 0.834 | 0.939 |
| RV Cardiac Output | 0.025 | -0.038, 0.088 | 0.433 | -0.057 | -0.125, 0.011 | 0.101 | 0.009 | -0.049, 0.067 | 0.759 | -0.046 | -0.109, 0.018 | 0.160 | 0.261 | 0.452 | 0.839 | 0.903 |
| LV Concentricity Index | -0.002 | -0.039, 0.036 | 0.924 | 0.047 | -0.007, 0.100 | 0.086 | 0.106 | 0.061, 0.151 | <0.001 | 0.079 | 0.018, 0.140 | 0.011 | 0.366 | 0.728 | 0.006 | 0.686 |
| LV Wall Thickness | 0.008 | -0.034, 0.051 | 0.698 | 0.015 | -0.041, 0.072 | 0.592 | 0.056 | 0.014, 0.099 | 0.009 | 0.032 | -0.021, 0.085 | 0.240 | 0.922 | 0.722 | 0.322 | 0.824 |
| LV Mass | 0.038 | 0.002, 0.073 | 0.036 | 0.013 | -0.029, 0.056 | 0.533 | 0.007 | -0.032, 0.047 | 0.713 | -0.011 | -0.057, 0.036 | 0.652 | 0.640 | 0.780 | 0.515 | 0.697 |

Results from fully adjusted models. β coefficients are reported per 1-SD increase in continuous exposures, with outcomes standardized. P-values were obtained from Wald tests and adjusted using the false discovery rate (FDR). [a]Premenopausal women vs Postmenopausal women; [b]Men < 50 years vs Men ≥ 50 years; [c]Premenopausal Women vs Men < 50 years; [d]Postmenopausal women vs Men ≥ 50 years. CI, confidence interval; LV, left ventricle; RV, right ventricle

**Supplementary Table 9.** Association of smoking status with cardiac parameters, and FDR-adjusted Wald test p-values for differences in regression coefficients.

| | | Women | | | | | | Men | | | | | | Wald Test | | | |
|---|---|---|---|---|---|---|---|---|---|---|---|---|---|---|---|---|---|
| | | Premenopausal | | | Postmenopausal | | | Age<50 | | | Age≥50 | | | | | | |
| **Outcome** | | **Beta** | **95% CI** | **p-value** | **Beta** | **95% CI** | **p-value** | **Beta** | **95% CI** | **p-value** | **Beta** | **95% CI** | **p-value** | **p-value[a]** | **p-value[b]** | **p-value[c]** | **p-value[d]** |
| LV Ejection Fraction | Former | -0.021 | -0.392, 0.350 | 0.912 | -0.068 | -0.516, 0.379 | 0.764 | 0.173 | -0.162, 0.508 | 0.310 | -0.248 | -0.652, 0.157 | 0.230 | 0.995 | 0.560 | 0.835 | 0.937 |
| | Current | -0.516 | -0.974, -0.058 | 0.027 | -0.315 | -0.947, 0.318 | 0.329 | -0.412 | -0.794, -0.030 | 0.035 | -0.739 | -1.323, -0.155 | 0.013 | 0.968 | 0.797 | 0.986 | 0.797 |
| LV End-diastolic Volume | Former | 0.583 | -0.022, 1.188 | 0.059 | 0.604 | -0.097, 1.305 | 0.091 | -0.493 | -1.141, 0.154 | 0.135 | 0.435 | -0.302, 1.172 | 0.247 | 0.999 | 0.472 | 0.222 | 0.986 |
| | Current | 0.502 | -0.245, 1.249 | 0.188 | 0.850 | -0.141, 1.841 | 0.093 | -0.137 | -0.876, 0.602 | 0.716 | 0.519 | -0.545, 1.584 | 0.339 | 0.947 | 0.797 | 0.727 | 0.982 |
| LV End-systolic Volume | Former | 0.237 | -0.065, 0.539 | 0.125 | 0.239 | -0.108, 0.587 | 0.177 | -0.264 | -0.586, 0.057 | 0.107 | 0.312 | -0.053, 0.677 | 0.094 | 0.999 | 0.222 | 0.234 | 0.986 |
| | Current | 0.479 | 0.106, 0.852 | 0.012 | 0.467 | -0.023, 0.958 | 0.062 | 0.238 | -0.128, 0.605 | 0.202 | 0.634 | 0.107, 1.162 | 0.018 | 0.999 | 0.722 | 0.797 | 0.982 |
| LV Stroke Volume | Former | 0.347 | -0.091, 0.784 | 0.120 | 0.365 | -0.133, 0.863 | 0.151 | -0.229 | -0.675, 0.216 | 0.313 | 0.123 | -0.378, 0.625 | 0.630 | 0.999 | 0.797 | 0.478 | 0.891 |
| | Current | 0.023 | -0.517, 0.563 | 0.933 | 0.382 | -0.321, 1.086 | 0.287 | -0.375 | -0.883, 0.133 | 0.147 | -0.115 | -0.839, 0.609 | 0.756 | 0.835 | 0.937 | 0.797 | 0.797 |
| LV Cardiac Output | Former | 31.014 | -8.452, 70.480 | 0.123 | 21.749 | -22.176, 65.674 | 0.332 | -10.570 | -47.628, 26.487 | 0.576 | 10.136 | -31.188, 51.459 | 0.631 | 0.986 | 0.843 | 0.560 | 0.986 |
| | Current | -21.079 | -69.847, 27.689 | 0.397 | 16.579 | -45.527, 78.685 | 0.601 | -23.680 | -65.957, 18.598 | 0.272 | -8.761 | -68.659, 51.137 | 0.774 | 0.797 | 0.986 | 0.999 | 0.937 |
| RV Ejection Fraction | Former | -0.305 | -0.815, 0.206 | 0.243 | -0.44 | -1.026, 0.145 | 0.140 | 0.454 | 0.019, 0.890 | 0.041 | 0.106 | -0.385, 0.598 | 0.671 | 0.986 | 0.797 | 0.234 | 0.597 |
| | Current | -0.190 | -0.821, 0.441 | 0.555 | -0.314 | -1.141, 0.514 | 0.458 | -0.197 | -0.693, 0.299 | 0.437 | 0.127 | -0.582, 0.837 | 0.725 | 0.986 | 0.843 | 0.999 | 0.835 |
| RV End-diastolic Volume | Former | 0.252 | -0.468, 0.972 | 0.493 | 0.334 | -0.453, 1.121 | 0.406 | -0.432 | -1.174, 0.309 | 0.253 | 0.303 | -0.524, 1.130 | 0.473 | 0.995 | 0.659 | 0.659 | 0.999 |
| | Current | 0.429 | -0.460, 1.317 | 0.344 | 0.326 | -0.787, 1.438 | 0.566 | -0.579 | -1.425, 0.266 | 0.179 | -0.936 | -2.131, 0.258 | 0.125 | 0.996 | 0.977 | 0.557 | 0.560 |
| RV End-systolic Volume | Former | 0.251 | -0.200, 0.702 | 0.276 | 0.417 | -0.031, 0.865 | 0.068 | -0.508 | -0.971, -0.045 | 0.031 | 0.061 | -0.420, 0.542 | 0.803 | 0.968 | 0.531 | 0.222 | 0.797 |
| | Current | 0.238 | -0.318, 0.795 | 0.401 | 0.306 | -0.326, 0.939 | 0.342 | -0.063 | -0.591, 0.464 | 0.814 | -0.485 | -1.180, 0.210 | 0.171 | 0.995 | 0.797 | 0.835 | 0.531 |
| RV Stroke Volume | Former | 0.001 | -0.498, 0.500 | 0.996 | -0.083 | -0.664, 0.497 | 0.778 | 0.076 | -0.413, 0.564 | 0.762 | 0.242 | -0.319, 0.802 | 0.398 | 0.986 | 0.982 | 0.986 | 0.835 |
| | Current | 0.190 | -0.426, 0.806 | 0.545 | 0.019 | -0.801, 0.839 | 0.963 | -0.516 | -1.073, 0.041 | 0.069 | -0.451 | -1.261, 0.359 | 0.275 | 0.986 | 0.999 | 0.531 | 0.835 |
| RV Cardiac Output | Former | 12.181 | -29.273, 53.634 | 0.565 | -6.365 | -53.496, 40.766 | 0.791 | 11.794 | -27.761, 51.349 | 0.559 | 18.749 | -26.144, 63.642 | 0.413 | 0.937 | 0.986 | 0.999 | 0.835 |
| | Current | -3.537 | -54.760, 47.687 | 0.892 | 0.384 | -66.255, 67.023 | 0.991 | -29.908 | -75.034, 15.218 | 0.194 | -29.966 | -95.038, 35.106 | 0.367 | 0.999 | 0.999 | 0.835 | 0.917 |

(continued)

**Supplementary Table 9.** Continued

| | | Women | | | | | | Men | | | | | | Wald test | | | |
|---|---|---|---|---|---|---|---|---|---|---|---|---|---|---|---|---|---|
| | | Premenopausal | | | Postmenopausal | | | Age<50 | | | Age≥50 | | | | | | |
| **Outcome** | | **Beta** | **95% CI** | **p-value** | **Beta** | **95% CI** | **p-value** | **Beta** | **95% CI** | **p-value** | **Beta** | **95% CI** | **p-value** | **p-value[a]** | **p-value[b]** | **p-value[c]** | **p-value[d]** |
| LV Concentricity Index | Former | -0.005 | -0.012, 0.001 | 0.112 | -0.007 | -0.017, 0.003 | 0.160 | 0.003 | -0.006, 0.011 | 0.540 | -0.005 | -0.017, 0.006 | 0.369 | 0.986 | 0.797 | 0.560 | 0.986 |
| | Current | 0.018 | 0.010, 0.026 | <0.001 | 0.028 | 0.014, 0.042 | <0.001 | 0.021 | 0.012, 0.031 | <0.001 | 0.028 | 0.012, 0.045 | <0.001 | 0.675 | 0.845 | 0.947 | 0.999 |
| LV Wall Thickness | Former | -0.010 | -0.027, 0.007 | 0.263 | -0.008 | -0.032, 0.016 | 0.492 | -0.013 | -0.031, 0.005 | 0.157 | -0.010 | -0.033, 0.013 | 0.412 | 0.999 | 0.986 | 0.986 | 0.999 |
| | Current | 0.063 | 0.042, 0.085 | <0.001 | 0.101 | 0.067, 0.135 | <0.001 | 0.052 | 0.031, 0.072 | <0.001 | 0.099 | 0.066, 0.133 | <0.001 | 0.472 | 0.222 | 0.835 | 0.999 |
| LV Mass | Former | 0.119 | -0.202, 0.441 | 0.467 | 0.214 | -0.190, 0.618 | 0.300 | -0.252 | -0.627, 0.124 | 0.190 | 0.195 | -0.258, 0.647 | 0.399 | 0.986 | 0.560 | 0.560 | 0.999 |
| | Current | 1.226 | 0.829, 1.623 | <0.001 | 2.055 | 1.484, 2.626 | <0.001 | 1.099 | 0.671, 1.528 | <0.001 | 2.144 | 1.490, 2.798 | <0.001 | 0.222 | 0.196 | 0.986 | 0.986 |

Results from fully adjusted models with additional adjustment for smoking exposure (pack-years). β coefficients are reported per 1-SD increase in continuous exposures, with outcomes standardized. P-values were obtained from Wald tests and adjusted using the false discovery rate (FDR).[a]Premenopausal women vs Postmenopausal women; [b]Men < 50 years vs Men ≥ 50 years; [c]Premenopausal Women vs Men < 50 years; [d]Postmenopausal women vs Men ≥ 50 years. CI, confidence interval; LV, left ventricle; RV, right ventricle

**Supplementary Table 10.** Association of alcohol use (units/day) with cardiac parameters, and FDR-adjusted Wald test p-values for differences in regression coefficients

| | Women | | | | | | Men | | | | | | Wald test | | | |
|---|---|---|---|---|---|---|---|---|---|---|---|---|---|---|---|---|
| | Premenopausal | | | Postmenopausal | | | Age<50 | | | Age≥50 | | | | | | |
| Outcome | Beta | 95% CI | p-value | Beta | 95% CI | p-value | Beta | 95% CI | p-value | Beta | 95% CI | p-value | p-value[a] | p-value[b] | p-value[c] | p-value[d] |
| LV Ejection Fraction | 0.018 | -0.008, 0.043 | 0.169 | 0.011 | -0.016, 0.037 | 0.438 | -0.014 | -0.028, 0.000 | 0.054 | 0.006 | -0.008, 0.020 | 0.395 | 0.839 | 0.183 | 0.146 | 0.875 |
| LV End-diastolic Volume | 0.008 | -0.013, 0.030 | 0.438 | 0.023 | 0.001, 0.044 | 0.042 | 0.012 | -0.002, 0.027 | 0.092 | -0.002 | -0.015, 0.011 | 0.807 | 0.613 | 0.381 | 0.875 | 0.226 |
| LV End-systolic Volume | -0.004 | -0.026, 0.018 | 0.717 | 0.010 | -0.012, 0.032 | 0.385 | 0.017 | 0.002, 0.031 | 0.024 | -0.004 | -0.017, 0.009 | 0.548 | 0.632 | 0.159 | 0.324 | 0.547 |
| LV Stroke Volume | 0.015 | -0.007, 0.038 | 0.187 | 0.026 | 0.003, 0.049 | 0.024 | 0.006 | -0.008, 0.021 | 0.410 | 0.001 | -0.013, 0.014 | 0.938 | 0.744 | 0.783 | 0.745 | 0.205 |
| LV Cardiac Output | 0.033 | 0.007, 0.058 | 0.012 | 0.023 | -0.002, 0.048 | 0.074 | 0.014 | -0.001, 0.029 | 0.061 | 0.004 | -0.009, 0.017 | 0.526 | 0.785 | 0.588 | 0.465 | 0.434 |
| RV Ejection Fraction | 0.003 | -0.023, 0.030 | 0.800 | 0.016 | -0.011, 0.043 | 0.259 | -0.009 | -0.024, 0.005 | 0.196 | 0.008 | -0.005, 0.021 | 0.216 | 0.759 | 0.244 | 0.655 | 0.803 |
| RV End-diastolic Volume | 0.007 | -0.015, 0.028 | 0.549 | 0.029 | 0.008, 0.050 | 0.006 | 0.010 | -0.004, 0.024 | 0.153 | -0.002 | -0.015, 0.010 | 0.728 | 0.360 | 0.431 | 0.885 | 0.082 |
| RV End-systolic Volume | 0.003 | -0.019, 0.025 | 0.778 | 0.013 | -0.006, 0.033 | 0.190 | 0.012 | -0.002, 0.027 | 0.095 | -0.007 | -0.019, 0.005 | 0.271 | 0.751 | 0.182 | 0.743 | 0.273 |
| RV Stroke Volume | 0.007 | -0.016, 0.031 | 0.543 | 0.034 | 0.009, 0.058 | 0.007 | 0.004 | -0.010, 0.019 | 0.558 | 0.003 | -0.010, 0.016 | 0.668 | 0.349 | 0.939 | 0.918 | 0.143 |
| RV Cardiac Output | 0.022 | -0.003, 0.047 | 0.090 | 0.030 | 0.004, 0.055 | 0.022 | 0.014 | -0.001, 0.029 | 0.070 | 0.006 | -0.008, 0.020 | 0.386 | 0.820 | 0.687 | 0.797 | 0.303 |
| LV Concentricity Index | 0.003 | -0.012, 0.018 | 0.707 | -0.006 | -0.026, 0.013 | 0.526 | 0.005 | -0.006, 0.017 | 0.374 | 0.010 | -0.003, 0.023 | 0.130 | 0.707 | 0.793 | 0.898 | 0.404 |
| LV Wall Thickness | -0.004 | -0.021, 0.013 | 0.647 | -0.014 | -0.035, 0.007 | 0.181 | 0.012 | 0.001, 0.024 | 0.028 | 0.004 | -0.007, 0.015 | 0.486 | 0.704 | 0.558 | 0.318 | 0.349 |
| LV Mass | 0.011 | -0.003, 0.026 | 0.112 | 0.017 | 0.001, 0.032 | 0.036 | 0.019 | 0.009, 0.029 | <0.001 | 0.012 | 0.002, 0.022 | 0.020 | 0.803 | 0.589 | 0.655 | 0.793 |

Results from fully adjusted models. β coefficients are reported per 1-SD increase in continuous exposures, with outcomes standardized. P-values were obtained from Wald tests and adjusted using the false discovery rate (FDR). [a]Premenopausal women vs Postmenopausal women; [b]Men < 50 years vs Men ≥ 50 years; [c]Premenopausal Women vs Men < 50 years; [d]Postmenopausal women vs Men ≥ 50 years. CI, confidence interval; LV, left ventricle; RV, right ventricle

**Supplementary Table 11.** Association of physical activity (per 10 MET-h/ week) with cardiac parameters, and FDR-adjusted Wald test p-values for differences in regression coefficients

| | Women | | | | | | Men | | | | | | Wald test | | | |
|---|---|---|---|---|---|---|---|---|---|---|---|---|---|---|---|---|
| | Premenopausal | | | Postmenopausal | | | Age<50 | | | Age≥50 | | | | | | |
| **Outcome** | **Beta** | **95% CI** | **p-value** | **Beta** | **95% CI** | **p-value** | **Beta** | **95% CI** | **p-value** | **Beta** | **95% CI** | **p-value** | **p-value[a]** | **p-value[b]** | **p-value[c]** | **p-value[d]** |
| LV Ejection Fraction | -0.001 | -0.008, 0.006 | 0.799 | 0.005 | -0.003, 0.013 | 0.196 | -0.004 | -0.010, 0.002 | 0.243 | 0.002 | -0.004, 0.009 | 0.503 | 0.510 | 0.434 | 0.782 | 0.780 |
| LV End-diastolic Volume | 0.018 | 0.012, 0.024 | <0.001 | 0.010 | 0.004, 0.016 | 0.002 | 0.032 | 0.026, 0.038 | <0.001 | 0.018 | 0.012, 0.024 | <0.001 | 0.244 | 0.023 | 0.023 | 0.249 |
| LV End-systolic Volume | 0.014 | 0.008, 0.020 | <0.001 | 0.005 | -0.002, 0.011 | 0.145 | 0.027 | 0.021, 0.033 | <0.001 | 0.013 | 0.007, 0.019 | <0.001 | 0.188 | 0.022 | 0.036 | 0.263 |
| LV Stroke Volume | 0.017 | 0.010, 0.023 | <0.001 | 0.011 | 0.005, 0.018 | <0.001 | 0.028 | 0.021, 0.034 | <0.001 | 0.018 | 0.011, 0.024 | <0.001 | 0.508 | 0.119 | 0.105 | 0.424 |
| LV Cardiac Output | 0.013 | 0.006, 0.020 | <0.001 | 0.010 | 0.003, 0.017 | 0.008 | 0.020 | 0.014, 0.027 | <0.001 | 0.013 | 0.007, 0.019 | <0.001 | 0.780 | 0.286 | 0.353 | 0.778 |
| RV Ejection Fraction | -0.009 | -0.017, -0.001 | 0.019 | -0.006 | -0.014, 0.002 | 0.154 | 0.002 | -0.004, 0.008 | 0.546 | -0.006 | -0.012, 0.000 | 0.057 | 0.780 | 0.253 | 0.138 | 0.998 |
| RV End-diastolic Volume | 0.022 | 0.015, 0.028 | <0.001 | 0.013 | 0.006, 0.019 | <0.001 | 0.035 | 0.029, 0.041 | <0.001 | 0.019 | 0.014, 0.025 | <0.001 | 0.163 | 0.005 | 0.023 | 0.317 |
| RV End-systolic Volume | 0.021 | 0.014, 0.027 | <0.001 | 0.012 | 0.007, 0.018 | <0.001 | 0.026 | 0.020, 0.032 | <0.001 | 0.019 | 0.013, 0.024 | <0.001 | 0.196 | 0.284 | 0.517 | 0.322 |
| RV Stroke Volume | 0.014 | 0.008, 0.021 | <0.001 | 0.008 | 0.001, 0.015 | 0.026 | 0.031 | 0.025, 0.037 | <0.001 | 0.013 | 0.007, 0.019 | <0.001 | 0.443 | <0.001 | 0.006 | 0.596 |
| RV Cardiac Output | 0.012 | 0.005, 0.019 | 0.001 | 0.008 | 0.000, 0.015 | 0.043 | 0.024 | 0.018, 0.031 | <0.001 | 0.009 | 0.003, 0.015 | 0.005 | 0.687 | 0.019 | 0.081 | 0.877 |
| LV Concentricity Index | 0.005 | 0.001, 0.009 | 0.020 | 0.002 | -0.003, 0.008 | 0.412 | 0.003 | -0.002, 0.008 | 0.255 | 0.002 | -0.004, 0.009 | 0.432 | 0.713 | 0.948 | 0.751 | 0.998 |
| LV Wall Thickness | 0.012 | 0.007, 0.017 | <0.001 | 0.008 | 0.002, 0.014 | 0.01 | 0.019 | 0.014, 0.023 | <0.001 | 0.013 | 0.007, 0.018 | <0.001 | 0.613 | 0.269 | 0.165 | 0.516 |
| LV Mass | 0.020 | 0.016, 0.024 | <0.001 | 0.012 | 0.007, 0.016 | <0.001 | 0.035 | 0.031, 0.039 | <0.001 | 0.022 | 0.018, 0.027 | <0.001 | 0.074 | 0.003 | <0.001 | 0.023 |

Results from fully adjusted models. β coefficients are reported per 1-SD increase in continuous exposures, with outcomes standardized. P-values were obtained from Wald tests and adjusted using the false discovery rate (FDR). [a]Premenopausal women vs Postmenopausal women; [b]Men < 50 years vs Men ≥ 50 years; [c]Premenopausal Women vs Men < 50 years; [d]Postmenopausal women vs Men ≥ 50 years. CI, confidence interval; LV, left ventricle; RV, right ventricle

**Supplementary Table 12.** Association of sleep duration shorter than 6.5 h (compared to 6.5-8.5h) with cardiac parameters, and FDR-adjusted Wald test p-values for differences in regression coefficients

| | Women | | | | | | Men | | | | | | Wald test (p-value) | | | |
|---|---|---|---|---|---|---|---|---|---|---|---|---|---|---|---|---|
| | Premenopausal | | | Postmenopausal | | | Age<50 | | | Age≥50 | | | | | | |
| **Outcome** | **Beta** | **95% CI** | **p-value** | **Beta** | **95% CI** | **p-value** | **Beta** | **95% CI** | **p-value** | **Beta** | **95% CI** | **p-value** | **p-value[a]** | **p-value[b]** | **p-value[c]** | **p-value[d]** |
| LV Ejection Fraction | -0.043 | -0.150, 0.063 | 0.426 | 0.101 | -0.037, 0.239 | 0.153 | -0.002 | -0.075, 0.072 | 0.961 | 0.016 | -0.081, 0.114 | 0.743 | 0.302 | 0.877 | 0.759 | 0.590 |
| LV End-diastolic Volume | 0.046 | -0.043, 0.136 | 0.311 | -0.062 | -0.175, 0.050 | 0.277 | -0.088 | -0.161, -0.014 | 0.019 | 0.044 | -0.049, 0.136 | 0.353 | 0.358 | 0.140 | 0.123 | 0.378 |
| LV End-systolic Volume | 0.057 | -0.036, 0.149 | 0.228 | -0.097 | -0.211, 0.017 | 0.096 | -0.073 | -0.148, 0.002 | 0.057 | 0.025 | -0.069, 0.118 | 0.608 | 0.165 | 0.315 | 0.148 | 0.303 |
| LV Stroke Volume | 0.028 | -0.068, 0.124 | 0.569 | -0.022 | -0.140, 0.096 | 0.709 | -0.078 | -0.152, -0.003 | 0.041 | 0.047 | -0.046, 0.140 | 0.321 | 0.751 | 0.165 | 0.272 | 0.619 |
| LV Cardiac Output | -0.010 | -0.118, 0.098 | 0.850 | -0.054 | -0.182, 0.074 | 0.407 | -0.039 | -0.116, 0.038 | 0.319 | 0.031 | -0.063, 0.126 | 0.515 | 0.798 | 0.510 | 0.820 | 0.547 |
| RV Ejection Fraction | 0.014 | -0.099, 0.127 | 0.807 | 0.005 | -0.134, 0.144 | 0.940 | -0.048 | -0.121, 0.026 | 0.204 | -0.104 | -0.195, -0.012 | 0.027 | 0.954 | 0.613 | 0.620 | 0.434 |
| RV End-diastolic Volume | 0.048 | -0.043, 0.139 | 0.301 | -0.076 | -0.183, 0.031 | 0.164 | -0.070 | -0.141, 0.002 | 0.058 | 0.053 | -0.035, 0.142 | 0.239 | 0.261 | 0.153 | 0.182 | 0.236 |
| RV End-systolic Volume | 0.024 | -0.070, 0.118 | 0.619 | -0.058 | -0.159, 0.043 | 0.258 | -0.027 | -0.101, 0.047 | 0.474 | 0.106 | 0.021, 0.190 | 0.015 | 0.500 | 0.116 | 0.656 | 0.097 |
| RV Stroke Volume | 0.054 | -0.047, 0.155 | 0.298 | -0.065 | -0.191, 0.060 | 0.310 | -0.085 | -0.160, -0.009 | 0.027 | -0.017 | -0.113, 0.079 | 0.728 | 0.373 | 0.527 | 0.146 | 0.778 |
| RV Cardiac Output | 0.008 | -0.099, 0.115 | 0.881 | -0.084 | -0.213, 0.046 | 0.205 | -0.046 | -0.123, 0.032 | 0.246 | -0.017 | -0.114, 0.080 | 0.733 | 0.541 | 0.809 | 0.680 | 0.673 |
| LV Concentricity Index | 0.013 | -0.051, 0.076 | 0.699 | 0.133 | 0.031, 0.234 | 0.010 | 0.097 | 0.037, 0.157 | 0.002 | 0.042 | -0.052, 0.135 | 0.381 | 0.187 | 0.592 | 0.220 | 0.431 |
| LV Wall Thickness | 0.077 | 0.004, 0.149 | 0.039 | 0.161 | 0.054, 0.268 | 0.003 | 0.107 | 0.050, 0.164 | <0.001 | 0.117 | 0.036, 0.198 | 0.005 | 0.436 | 0.921 | 0.751 | 0.751 |
| LV Mass | 0.058 | -0.002, 0.118 | 0.057 | 0.068 | -0.012, 0.149 | 0.096 | -0.004 | -0.057, 0.050 | 0.890 | 0.093 | 0.022, 0.163 | 0.010 | 0.921 | 0.148 | 0.345 | 0.813 |

Results from fully adjusted models. β coefficients are reported per 1-SD increase in continuous exposures, with outcomes standardized. P-values were obtained from Wald tests and adjusted using the false discovery rate (FDR). [a]Premenopausal women vs Postmenopausal women; [b]Men < 50 years vs Men ≥ 50 years; [c]Premenopausal Women vs Men < 50 years; [d]Postmenopausal women vs Men ≥ 50 years. CI, confidence interval; LV, left ventricle; RV, right ventricle

**Supplementary Table 13.** Association of sleep duration longer than 8.5 h (compared to 6.5-8.5h) with cardiac parameters, and FDR-adjusted Wald test p-values for differences in regression coefficients

| | Women | | | | | | Men | | | | | | Wald test (p-value) | | | |
|---|---|---|---|---|---|---|---|---|---|---|---|---|---|---|---|---|
| | Premenopausal | | | Postmenopausal | | | Age<50 | | | Age≥50 | | | | | | |
| **Outcome** | **Beta** | **95% CI** | **p-value** | **Beta** | **95% CI** | **p-value** | **Beta** | **95% CI** | **p-value** | **Beta** | **95% CI** | **p-value** | **p-value[a]** | **p-value[b]** | **p-value[c]** | **p-value[d]** |
| LV Ejection Fraction | -0.055 | -0.122, 0.012 | 0.107 | -0.082 | -0.158, -0.007 | 0.032 | -0.046 | -0.111, 0.020 | 0.172 | -0.092 | -0.172, -0.012 | 0.024 | 0.793 | 0.630 | 0.921 | 0.928 |
| LV End-diastolic Volume | -0.040 | -0.097, 0.016 | 0.164 | -0.092 | -0.153, -0.031 | 0.003 | -0.059 | -0.125, 0.006 | 0.074 | -0.094 | -0.169, -0.019 | 0.014 | 0.461 | 0.743 | 0.813 | 0.990 |
| LV End-systolic Volume | 0.006 | -0.052, 0.064 | 0.844 | -0.027 | -0.090, 0.035 | 0.391 | -0.024 | -0.090, 0.043 | 0.486 | -0.014 | -0.091, 0.062 | 0.712 | 0.692 | 0.927 | 0.751 | 0.896 |
| LV Stroke Volume | -0.064 | -0.124, -0.003 | 0.040 | -0.117 | -0.181, -0.053 | <0.001 | -0.071 | -0.137, -0.005 | 0.036 | -0.129 | -0.204, -0.053 | <0.001 | 0.488 | 0.515 | 0.935 | 0.909 |
| LV Cardiac Output | -0.053 | -0.121, 0.015 | 0.125 | -0.050 | -0.120, 0.020 | 0.160 | -0.062 | -0.130, 0.007 | 0.077 | -0.111 | -0.188, -0.034 | 0.005 | 0.972 | 0.612 | 0.928 | 0.506 |
| RV Ejection Fraction | -0.039 | -0.110, 0.032 | 0.281 | 0.037 | -0.039, 0.112 | 0.344 | -0.035 | -0.100, 0.031 | 0.299 | 0.012 | -0.062, 0.087 | 0.748 | 0.378 | 0.613 | 0.957 | 0.810 |
| RV End-diastolic Volume | -0.068 | -0.126, -0.011 | 0.020 | -0.133 | -0.192, -0.075 | <0.001 | -0.063 | -0.127, 0.001 | 0.053 | -0.096 | -0.169, -0.024 | 0.009 | 0.324 | 0.743 | 0.947 | 0.686 |
| RV End-systolic Volume | -0.022 | -0.081, 0.038 | 0.471 | -0.110 | -0.165, -0.055 | <0.001 | -0.032 | -0.098, 0.034 | 0.341 | -0.077 | -0.146, -0.008 | 0.029 | 0.148 | 0.613 | 0.909 | 0.707 |
| RV Stroke Volume | -0.088 | -0.151, -0.024 | 0.007 | -0.106 | -0.175, -0.038 | 0.002 | -0.070 | -0.136, -0.003 | 0.042 | -0.079 | -0.157, -0.001 | 0.046 | 0.834 | 0.925 | 0.834 | 0.800 |
| RV Cardiac Output | -0.082 | -0.149, -0.015 | 0.017 | -0.053 | -0.123, 0.018 | 0.142 | -0.070 | -0.140, -0.001 | 0.046 | -0.076 | -0.155, 0.003 | 0.059 | 0.780 | 0.949 | 0.903 | 0.819 |
| LV Concentricity Index | 0.015 | -0.026, 0.055 | 0.478 | 0.051 | -0.005, 0.106 | 0.073 | 0.061 | 0.007, 0.114 | 0.027 | 0.087 | 0.011, 0.163 | 0.025 | 0.558 | 0.785 | 0.411 | 0.697 |
| LV Wall Thickness | -0.026 | -0.072, 0.019 | 0.259 | 0.022 | -0.037, 0.080 | 0.463 | 0.064 | 0.013, 0.115 | 0.013 | 0.015 | -0.051, 0.081 | 0.647 | 0.436 | 0.506 | 0.074 | 0.939 |
| LV Mass | -0.026 | -0.064, 0.012 | 0.177 | -0.045 | -0.089, -0.001 | 0.045 | -0.004 | -0.051, 0.044 | 0.873 | -0.058 | -0.116, -0.000 | 0.049 | 0.751 | 0.381 | 0.715 | 0.845 |

Results from fully adjusted models. β coefficients are reported per 1-SD increase in continuous exposures, with outcomes standardized. P-values were obtained from Wald tests and adjusted using the false discovery rate (FDR). [a]Premenopausal women vs Postmenopausal women; [b]Men < 50 years vs Men ≥ 50 years; [c]Premenopausal Women vs Men < 50 years; [d]Postmenopausal women vs Men ≥ 50 years. CI, confidence interval; LV, left ventricle; RV, right ventricle

**Supplementary Table 14.** Association of sleep problems with cardiac parameters, and FDR-adjusted Wald test p-values for differences in regression coefficients

| | Women | | | | | | Men | | | | | | Wald test | | | |
|---|---|---|---|---|---|---|---|---|---|---|---|---|---|---|---|---|
| | Premenopausal | | | Postmenopausal | | | Age<50 | | | Age≥50 | | | | | | |
| **Outcome** | **Beta** | **95% CI** | **p-value** | **Beta** | **95% CI** | **p-value** | **Beta** | **95% CI** | **p-value** | **Beta** | **95% CI** | **p-value** | **p-value[a]** | **p-value[b]** | **p-value[c]** | **p-value[d]** |
| LV Ejection Fraction | -0.039 | -0.098, 0.020 | 0.197 | 0.003 | -0.068, 0.074 | 0.939 | -0.038 | -0.092, 0.016 | 0.171 | -0.019 | -0.094, 0.056 | 0.620 | 0.630 | 0.830 | 0.991 | 0.825 |
| LV End-diastolic Volume | -0.003 | -0.053, 0.046 | 0.897 | 0.043 | -0.015, 0.101 | 0.143 | -0.093 | -0.147, -0.040 | <0.001 | -0.105 | -0.175, -0.034 | 0.004 | 0.481 | 0.899 | 0.099 | 0.023 |
| LV End-systolic Volume | 0.021 | -0.030, 0.072 | 0.414 | 0.032 | -0.027, 0.091 | 0.287 | -0.048 | -0.103, 0.007 | 0.085 | -0.071 | -0.143, 0.001 | 0.052 | 0.888 | 0.803 | 0.237 | 0.140 |
| LV Stroke Volume | -0.020 | -0.073, 0.033 | 0.458 | 0.041 | -0.020, 0.101 | 0.187 | -0.104 | -0.158, -0.049 | <0.001 | -0.104 | -0.175, -0.033 | 0.004 | 0.358 | 0.998 | 0.146 | 0.031 |
| LV Cardiac Output | 0.012 | -0.047, 0.072 | 0.680 | 0.036 | -0.030, 0.102 | 0.281 | -0.086 | -0.143, -0.030 | 0.003 | -0.087 | -0.159, -0.015 | 0.018 | 0.793 | 0.998 | 0.107 | 0.090 |
| RV Ejection Fraction | 0.017 | -0.046, 0.079 | 0.603 | 0.002 | -0.069, 0.074 | 0.954 | -0.042 | -0.096, 0.012 | 0.129 | -0.016 | -0.086, 0.054 | 0.652 | 0.875 | 0.782 | 0.392 | 0.845 |
| RV End-diastolic Volume | -0.020 | -0.070, 0.030 | 0.438 | 0.016 | -0.039, 0.072 | 0.559 | -0.102 | -0.155, -0.049 | <0.001 | -0.086 | -0.154, -0.018 | 0.013 | 0.602 | 0.845 | 0.138 | 0.116 |
| RV End-systolic Volume | -0.029 | -0.081, 0.023 | 0.272 | 0.011 | -0.041, 0.062 | 0.688 | -0.052 | -0.107, 0.002 | 0.059 | -0.055 | -0.120, 0.010 | 0.096 | 0.544 | 0.971 | 0.775 | 0.325 |
| RV Stroke Volume | -0.004 | -0.059, 0.052 | 0.899 | 0.016 | -0.049, 0.080 | 0.629 | -0.112 | -0.167, -0.056 | <0.001 | -0.084 | -0.157, -0.011 | 0.024 | 0.810 | 0.780 | 0.061 | 0.176 |
| RV Cardiac Output | 0.024 | -0.035, 0.083 | 0.420 | 0.017 | -0.050, 0.083 | 0.627 | -0.098 | -0.155, -0.041 | <0.001 | -0.071 | -0.145, 0.003 | 0.061 | 0.928 | 0.782 | 0.038 | 0.264 |
| LV Concentricity Index | 0.005 | -0.031, 0.040 | 0.802 | -0.041 | -0.093, 0.011 | 0.125 | 0.055 | 0.011, 0.099 | 0.015 | 0.075 | 0.004, 0.147 | 0.039 | 0.381 | 0.809 | 0.254 | 0.075 |
| LV Wall Thickness | 0.015 | -0.025, 0.055 | 0.475 | -0.011 | -0.066, 0.044 | 0.702 | 0.049 | 0.007, 0.090 | 0.023 | 0.062 | 0.000, 0.124 | 0.050 | 0.707 | 0.845 | 0.505 | 0.263 |
| LV Mass | -0.004 | -0.037, 0.029 | 0.823 | 0.007 | -0.035, 0.048 | 0.755 | -0.034 | -0.073, 0.005 | 0.086 | -0.050 | -0.104, 0.004 | 0.071 | 0.834 | 0.809 | 0.500 | 0.301 |

Results from fully adjusted models. β coefficients are reported per 1-SD increase in continuous exposures, with outcomes standardized. P-values were obtained from Wald tests and adjusted using the false discovery rate (FDR). [a]Premenopausal women vs Postmenopausal women; [b]Men < 50 years vs Men ≥ 50 years; [c]Premenopausal Women vs Men < 50 years; [d]Postmenopausal women vs Men ≥ 50 years. CI, confidence interval; LV, left ventricle; RV, right ventricle
